# Assessing innovative care models for musculoskeletal disorders’ management in the emergency department using Time-Driven Activity-Based Costing

**DOI:** 10.1101/2024.08.14.24311988

**Authors:** Rose Gagnon, Kadija Perreault, Jason R. Guertin, Luc J. Hébert, Simon Berthelot

**Author notes:** **Corresponding Author:** Simon Berthelot, MD, MSc Population Health and Optimal Health Practices Axis, CHU de Québec – Université Laval Research Centre 2705, Boulevard Laurier, R-00765 Quebec City (QC), Canada G1V 4G2.

## Abstract

**Objectives:** Compare the average cost of an emergency department (ED) visit between three ED care models, namely management by an emergency physician (EP) alone (usual care), management by a primary contact physiotherapist (PT) and an EP (intervention), and management by a PT alone (sensitivity analysis).

**Methods:** Cost study (Canadian Public Payer perspective) based on data collected during a pragmatic randomized clinical trial (2018-2019) conducted in an urban Canadian academic ED (CHUL, Quebec City, Canada; n=78, 18-80 years old). Costs incurred for the management of persons presenting to the ED for a minor musculoskeletal disorder (MSKD) were calculated using Time-Driven Activity-Based Costing, in which time invested with a patient determines care costs. The main outcome measure was the average cost of an ED visit. Generalized linear models with Gamma distributions and log links were used to assess whether there were significant differences in average costs between the care models.

**Results:** Mean ED visit cost was $267.08 (2019 $CAD, 95%CI: $212.75, $346.40) for PT and EP management, compared with $245.14 for EP management ($169.46, $336.72), resulting in a non-significant absolute difference of 21.94 CAD/patient ($-87.33, $132.63) between models (p=.60). Sensitivity analyses showed that the average cost of ED management by a PT was $194.38 ($161.50, $234.34), representing a non-significant average saving of 50.76 CAD/patient ($- 156.91, $37.54) compared to EP management.

**Conclusion:** This study is a first step towards a better understanding of the costs incurred by the Canadian Public Payer for the management of persons presenting with MSKDs in the ED. Primary contact physiotherapists have the potential to complement care of MSKD ED patients without increasing healthcare costs.

## Introduction

Emergency department (ED) overcrowding has a number of deleterious consequences, such as reduced quality of care, avoidable medical errors, depletion of available resources, and job dissatisfaction among ED care providers.^1–6^ To categorize the causes of overcrowding, ED patient flow can be divided in three distinct phases: *input*, *throughput*, and *output*.^7^ The *input* phase is characterized by all events, conditions or characteristics of the healthcare system that influence the demand for emergency care.^7^ The *throughput* phase includes all the processes happening during the ED visit.^7^ The *output* phase refers to all factors affecting the ability of ED staff to move their patients out of the ED by either admitting or discharging them (e.g., lack of healthcare resources in the community, blocked access to hospital beds).^7,8^

Potential strategies to improve the *throughput* phase include the integration of physiotherapists in the ED to manage persons presenting with minor musculoskeletal disorders (MSKDs)^9^ immediately after triage by the nurse. While the exact titles taken by physiotherapists working in EDs have varied between studies, have evolved over time, and may vary internationally depending on regulations (e.g., primary contact physiotherapist,^10,11^ advanced practice physiotherapist,^12,13^ extended scope physiotherapist^14^), the roles of these physiotherapists usually include independent assessment, diagnosis, management, and discharge, and may include the autonomy to perform some medical delegated acts, often referred to as advanced practice models of care by several professional associations or regulatory physiotherapy colleges.^15^ Until now, such an integration of physiotherapists in EDs has been associated with a number of benefits, such as a reduction in time to care,^12,16–19^ length of stay in the ED,^12,16,17,19,20^ consultations with various healthcare professionals,^21^ and prescription of medication^21,22^ (including opioids^20^) and imaging tests^18,20,21,23^. Previous work by our team showed that integrating physiotherapists in the ED is also associated with a reduction in the use of healthcare system services and resources for up to three months following the initial ED visit (i.e., new ED visits for the same condition, use of medication) when compared to usual emergency physician care.^22^ However, very few studies have looked at the costs associated with such an ED care model,^24^ mainly because there is little or no information available on the costs of ED care processes used to manage persons presenting with minor MSKDs.

Several cost measurement methods have been used over the years in an attempt to measure costs specific to the healthcare system. The most frequently used methods include, but are not limited to, cost-to-charge ratios, relative value units and activity-based costing.^25^ These methods have a number of methodological limitations, in that they often are based on subjective measures^26^ and/or on the amount reimbursed for a procedure or health service.^25,27,28^ Moreover, the implementation of these costing methods is generally very expensive and time-consuming,^28^ resulting in cost models that are rarely updated which leads to outdated cost estimates.^26,29^ One of the costing methods developed to overcome these methodological limitations is Time-Driven Activity Based Costing (TDABC). Unlike previous methods, it allows to obtain a treatment cost per patient specific to a health condition.^27^ TDABC also allows for variability 1) in the clinical presentation of patients with the same health condition, and 2) in the different declinations of care processes (e.g., ankle vs. lumbar X-ray).^28^ This adaptability makes it an ideal costing method for changing environments such as the ED, where a wide variety of care processes are used.^25,29^ This method has indeed recently been adapted and validated for use in EDs for a variety of minor disorders (e.g., upper respiratory tract infections, abdominal pain).^30^

Therefore, the aim of this study was to compare, using the TDABC method, the average cost of an ED visit for a minor MSKD under the management of a primary contact physiotherapist and an emergency physician with that of usual management by an emergency physician.

## Methods

### Study design

Cost study performed using TDABC and based on a secondary analysis of cost data collected during a pilot pragmatic randomized clinical trial (RCT). This trial was conducted between September 1^st^, 2018, and March 31^st^, 2019, at the CHU de Québec – Université Laval, the third largest healthcare institution in Canada (*Centre hospitalier de l’Université Laval* (CHUL), Quebec City, Canada). As Canada provides a universal healthcare system, expenses incurred by patients when presenting to the hospital, including the ED, are entirely covered by the government. Therefore, our cost study was carried out according to a Public Payer’s Perspective and included all healthcare expenses incurred during the ED visit (including those related to physician remuneration who are paid by a public governmental third-party payer, the *Régie de l’assurance maladie du Québec*). Further details on the use of cost data collected during the trial can be found in our published protocol.^31^ The present project was approved by the CHU de Québec - Université Laval’s Research Ethics Committee (#MP-20-2019-4307).

The RCT was designed to compare the effects of two care models in the ED, i.e. 1) usual management by an emergency physician (control group); and 2) management by a primary contact physiotherapist and an emergency physician (intervention group). To be included in the trial, participants (n=78) had to meet the criteria shown in Figure 1:

**Figure 1.**
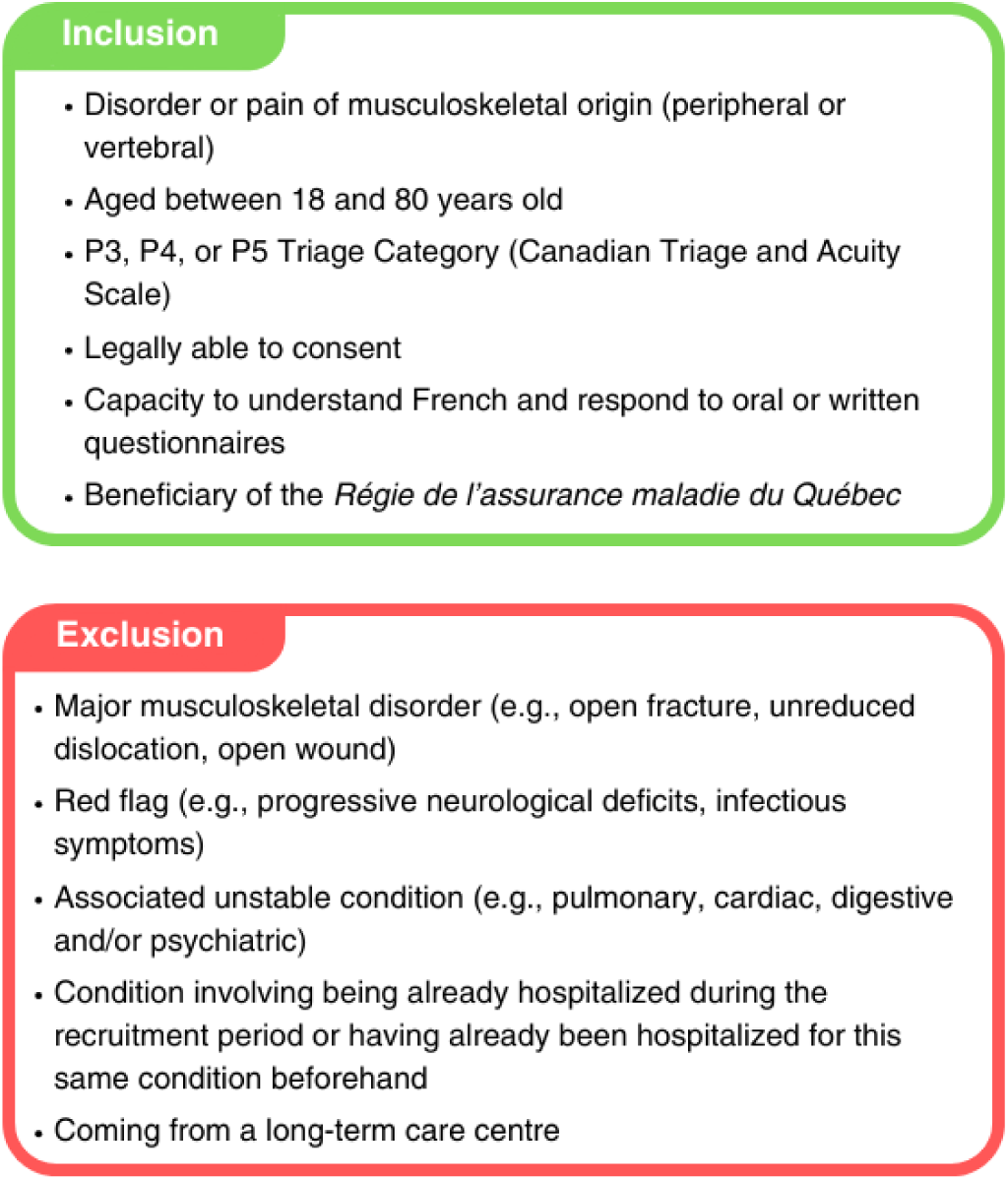
Inclusion and exclusion criteria used during the pilot pragmatic randomized clinical trial

Once recruited, persons presenting with a minor MSKD were randomized into the two different groups. In the control group, given the pragmatic nature of the clinical trial, the emergency physician was free to use any intervention deemed necessary during the ED visit. The second group (intervention) was managed by a primary contact physiotherapist and an emergency physician. The participants were first seen by a physiotherapist, who carried out their subjective and objective examinations. The physiotherapist was free to recommend any intervention deemed appropriate. After the evaluation, participants returned to the waiting room and were seen by the emergency physician. The emergency physician was encouraged to discuss patient management with the primary contact physiotherapist but was free to prescribe any intervention deemed appropriate. At the time of the clinical trial, physiotherapists could not discharge patients autonomously in the host setting because of regulatory local bylaws. Participants in both groups were therefore allowed to leave the ED once the emergency physician had recommended discharge. More information on the interventions performed and the course of management in the ED can be found in our previously published article.^22^

### Costing approach

TDABC was used to calculate the average cost of an ED visit in both groups. This method bases its cost estimates on the time spent providing care to the patient. As originally introduced by Kaplan and Porter, ^25,27^ TDABC can be broken down into seven main steps, presented in Box 1:

**Box 1.**
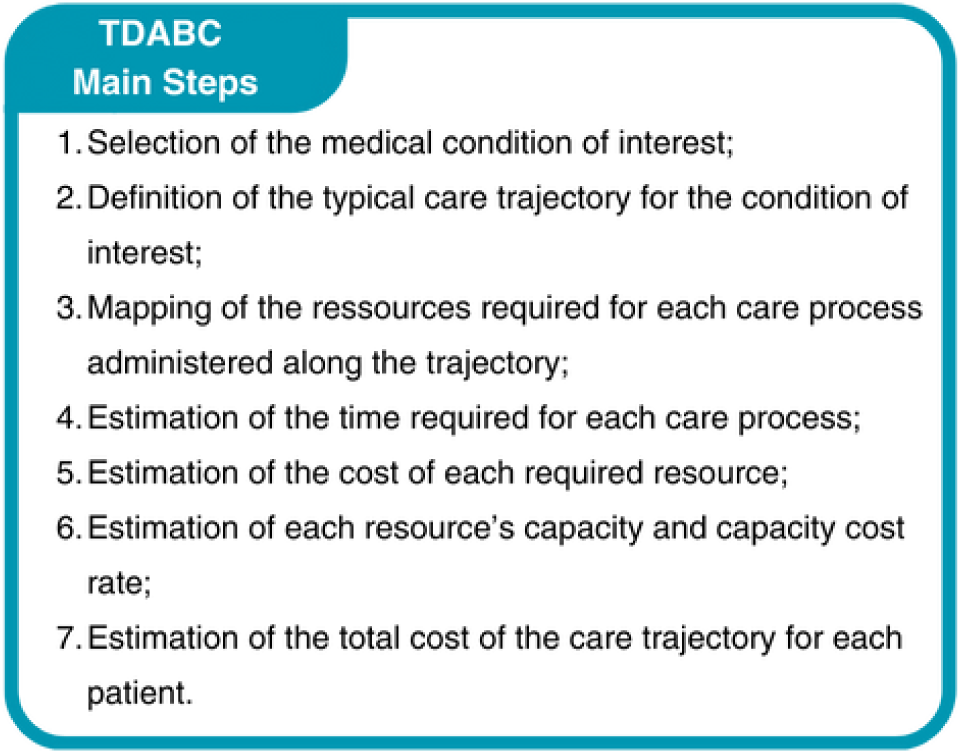
Main steps of the Time-Driven Activity-Based Costing Method

As the use of TDABC in the context of our trial has already been discussed,^31^ the steps below will focus on the various methodological choices made during our application of the method.

#### 1. Selection of the medical condition of interest

All participants recruited during the RCT were included in the cost study (n=78, 40 in the intervention group and 38 in the control group). Some (n=6) of the participants left the ED without being seen by a physician. They were nevertheless included, since a visit of this type contributes to the financial burden of an ED (e.g., human resources, stretchers).

#### 2. Definition of the typical care trajectory for the condition of interest

Each of the participants recruited during the RCT followed an ED care trajectory summarized in Figure 2:

**Figure 2.**
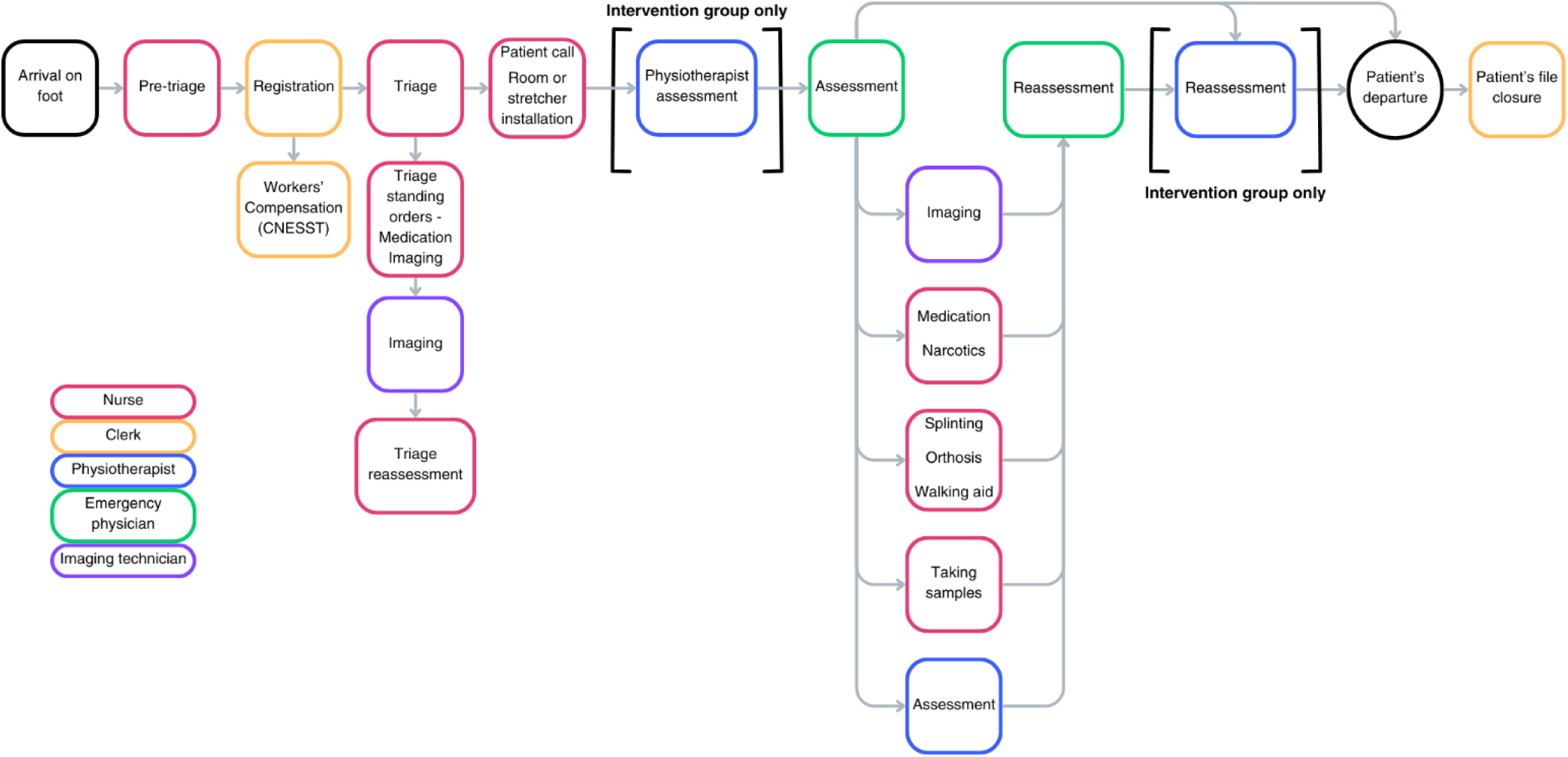
Typical emergency department care trajectory of a patient presenting for a minor musculoskeletal disorder

Only participants in the intervention group were managed by a physiotherapist following triage by the nurse. In addition, while Figure 2 shows all the possible interventions (care processes) received by persons presenting with a minor MSKD in the ED, participants may not have received all of them, as some interventions might not have been considered useful during their visit.

a. 3. *Mapping of the resources required for each intervention (i.e., care process)*

This step was carried out as part of work previously published by Berthelot et al.; further details on the method used to carry out the mapping can be found in another publication.^30^ Detailed mapping of the required resources for each ED care process can be found in Table S1.

Once all care processes were mapped, a member of the research team (RG) reviewed each study participant’s electronic medical record to compile all the care processes administered during their ED visit. This approach enabled the research team to accurately reconstruct the specific care pathway of each participant included in the study.

#### 4. Estimation of the time required for each care process

The durations of the most frequent ED care processes were measured using a time-motion software (more details here ^30^). Durations of less frequent care processes were estimated by surveying healthcare professionals involved in administering these processes through short interviews or an online survey questionnaire. The durations of all care processes identified during the mapping process, as well as further details on their measurement process (direct measurement vs. estimation) can be found in Table S2.

#### 5. Estimation of the cost of each required resource

Unless otherwise specified, all data required to estimate costs of each required resource were extracted from the 2018-2019 fiscal year expense reports provided by the CHU de Québec - Université Laval Finance Department.

### Human resources

Wages considered for all types of human resources employed in the ED (i.e., nurse, clerk, nursing assistant, imaging technician, physiotherapist, social worker, occupational therapist, hospital porter) consisted of salary, benefits and bonuses paid during the fiscal year 2018-2019.

### Physicians

Physicians are independent workers paid by the Public Payer, but their income is not accounted for in a hospital’s financial data. Therefore, we estimated the total cost associated with this resource as follows:
1. The total number of shifts performed during the 2018-2019 fiscal year in the included ED was extracted from the official physicians’ work schedule.
2. Shifts performed were counted into two categories, based on whether they were performed by an emergency physician who was a general practitioner or an emergency medicine specialist. This was used to derive the proportion of ED shifts attributable to each specialty.
3. Average annual salaries for each specialty were obtained from publicly available ministerial data.
4. The average annual wage for an ED physician was calculated using the following equation: 

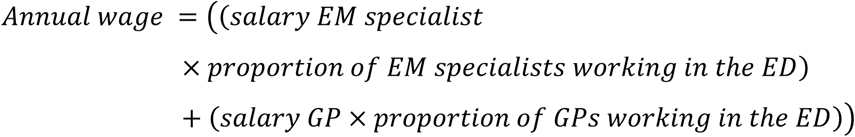 Where EM: emergency medicine and GP: general practitioner
5. Finally, the total amount of physicians’ wages for the study period was estimated by multiplying the average annual wage of an ED physician by the number of full-time equivalents practicing in the ED.

As for physician fees related to imaging tests and/or ECGs performed, a fixed fee corresponding to the remuneration offered for the service performed was added to the cost of performing the test in the ED, to account for the cost of interpretation by a specialist (i.e., radiologist, cardiologist).

Material resources (i.e., radiology equipment)

The annual cost of each imaging device (i.e., CT scan, MRI, X-ray, ultrasound) was calculated using the following equation:

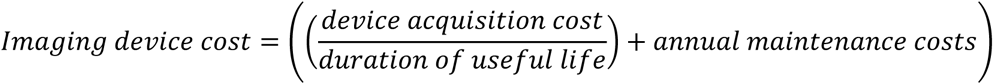

### Consumables

Expenses related to consumables were accounted for by hospital department based on the human resources involved in ED care. They were extracted from the expense reports provided to the CHU de Québec - Université Laval by each of the concerned departments for the 2018-2019 fiscal year.

#### Traceable supplies

The use of certain specific consumables (i.e. drugs, orthoses, walking aids and laboratory tests) could be traced directly via the ED’s electronic medical record. The unit cost of each traceable supply used by a patient was directly added to the cost of his or her ED visit. The following data sources were used for each type of traceable supply:

a. Drugs: drug supply contract negotiated by the CHU de Québec – Université Laval
b. Orthoses and walking aids: negotiated supply contract with a private supplier
c. Laboratory tests: publicly available ministerial data

### Overhead

Unlike the above-mentioned resources, data used to calculate overhead were retrieved from public reports issued by the CHU de Québec – Université Laval. A detailed description of the method used by our team to calculate overhead, and the budget items considered can be found in the Methods Supplement.

#### 6. Estimation of each resource’s capacity and capacity cost rate

The capacity cost rate of the various resources involved in ED care and used to calculate the average ED visit cost per care model are reported in Table 1. The methods used to estimate the capacity (i.e., time available for direct patient care in minutes) and the capacity cost rate (in $/minute) for each resource are detailed below.

**Table 1.**
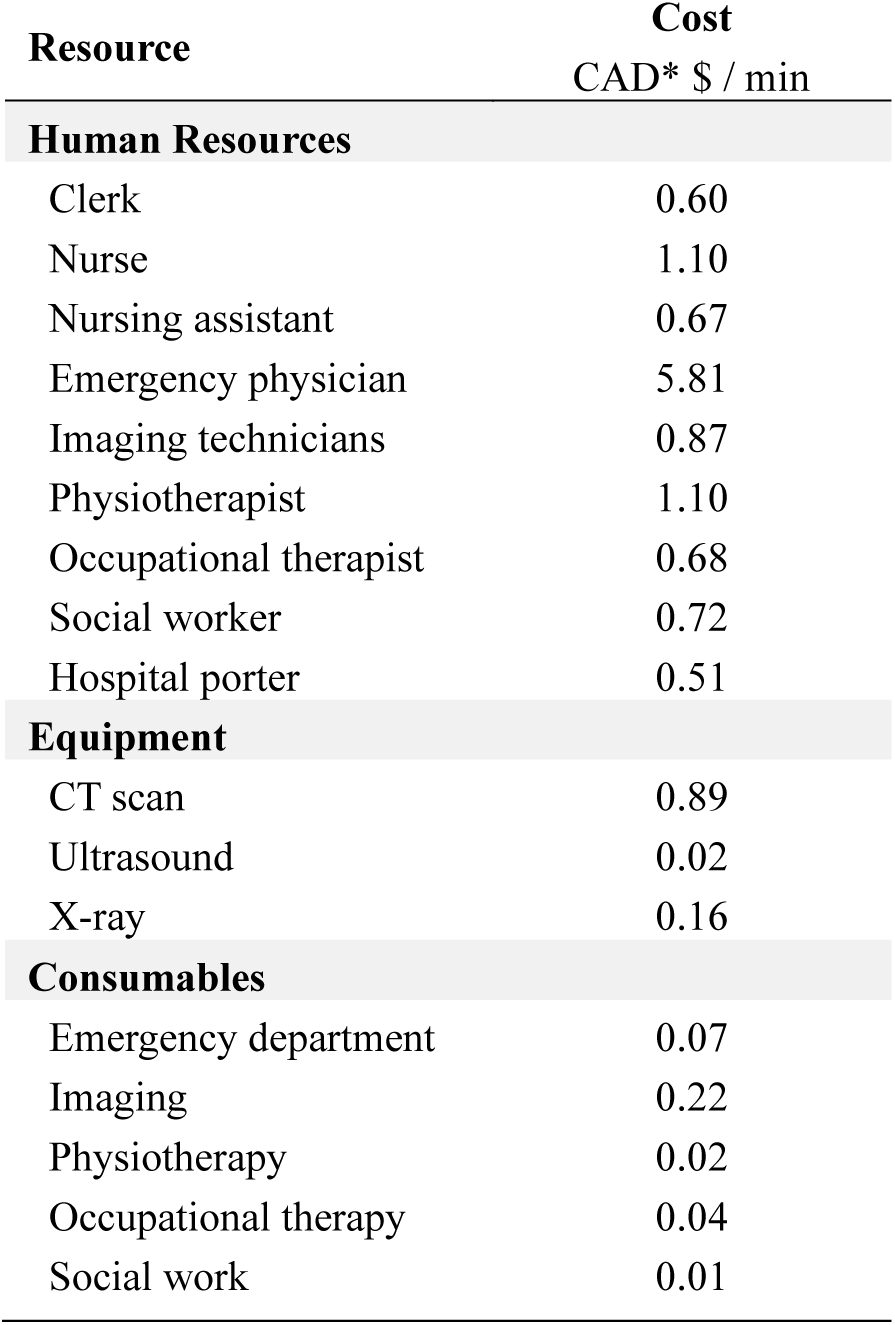
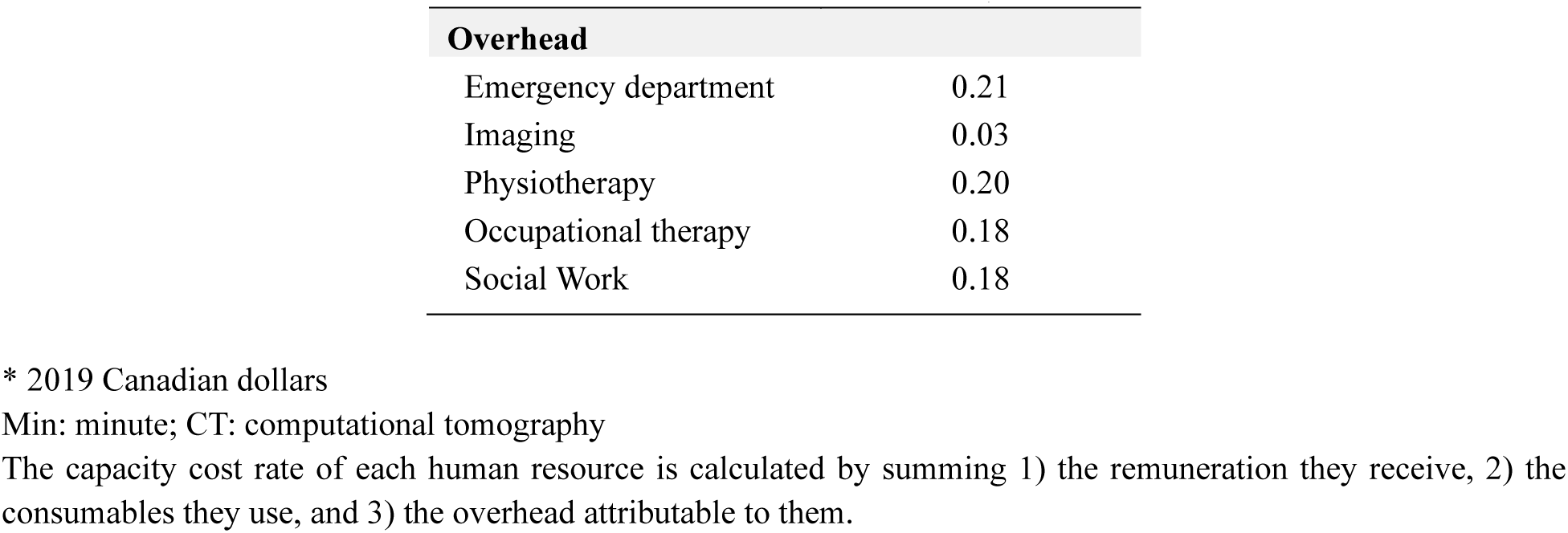
Capacity cost rate of emergency department resources.

### Human resources

The capacity of each human resource was estimated using the total number of hours worked in patient care (excluding trainings and meetings) during the study period. Seven percent of hours worked were subtracted to account for non-productivity (e.g., breaks).

### Emergency physicians

The total number of hours worked in patient care over the year by the emergency physicians was calculated from the official shift schedule. Seven percent of the hours worked were subtracted for the same reasons as above.

Material resources (i.e., radiology equipment)

The number of minutes of ED use for each imaging device was extracted from the annual utilization report provided by the CHU de Québec – Université Laval to the provincial Ministry of Health and Social Services.

### Consumables

The total consumable capacity was recorded by clinical department involved in ED care and corresponded to the time available for direct patient care of each department.

### Capacity cost rate calculation

The capacity cost rate for each resource was obtained using the following equation:

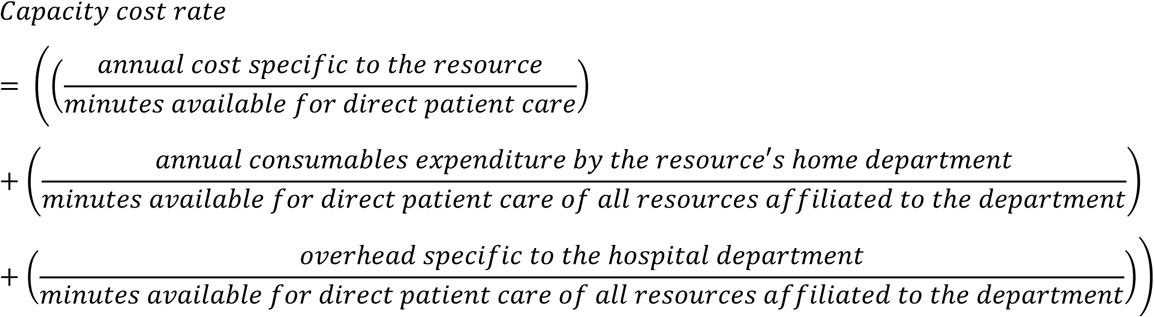

#### 7. Estimation of the total cost of the ED visit for each patient

The unit cost of each ED care process was obtained in two steps. First, the capacity cost rate of each resource required to carry out a care process (obtained through Step #6) was multiplied by the duration in minutes of the process (Table S2, obtained through Step #4). The costs associated with each necessary resource were then added together to obtain the total cost of carrying out the care process of interest. The unit costs associated with each of the care processes mapped in the ED and used to calculate the total cost of each ED visit are presented in Table 2:

**Table 2.**
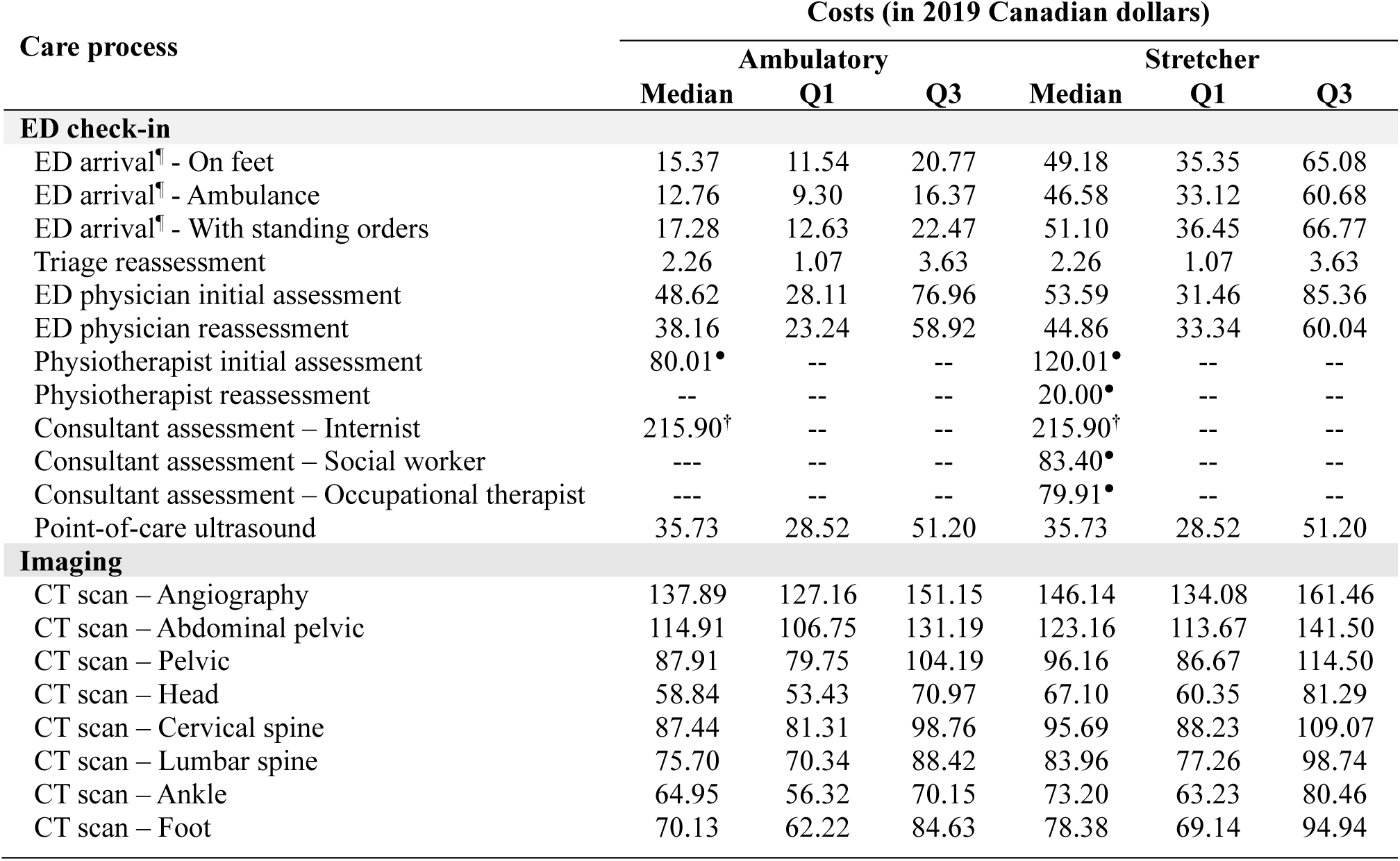
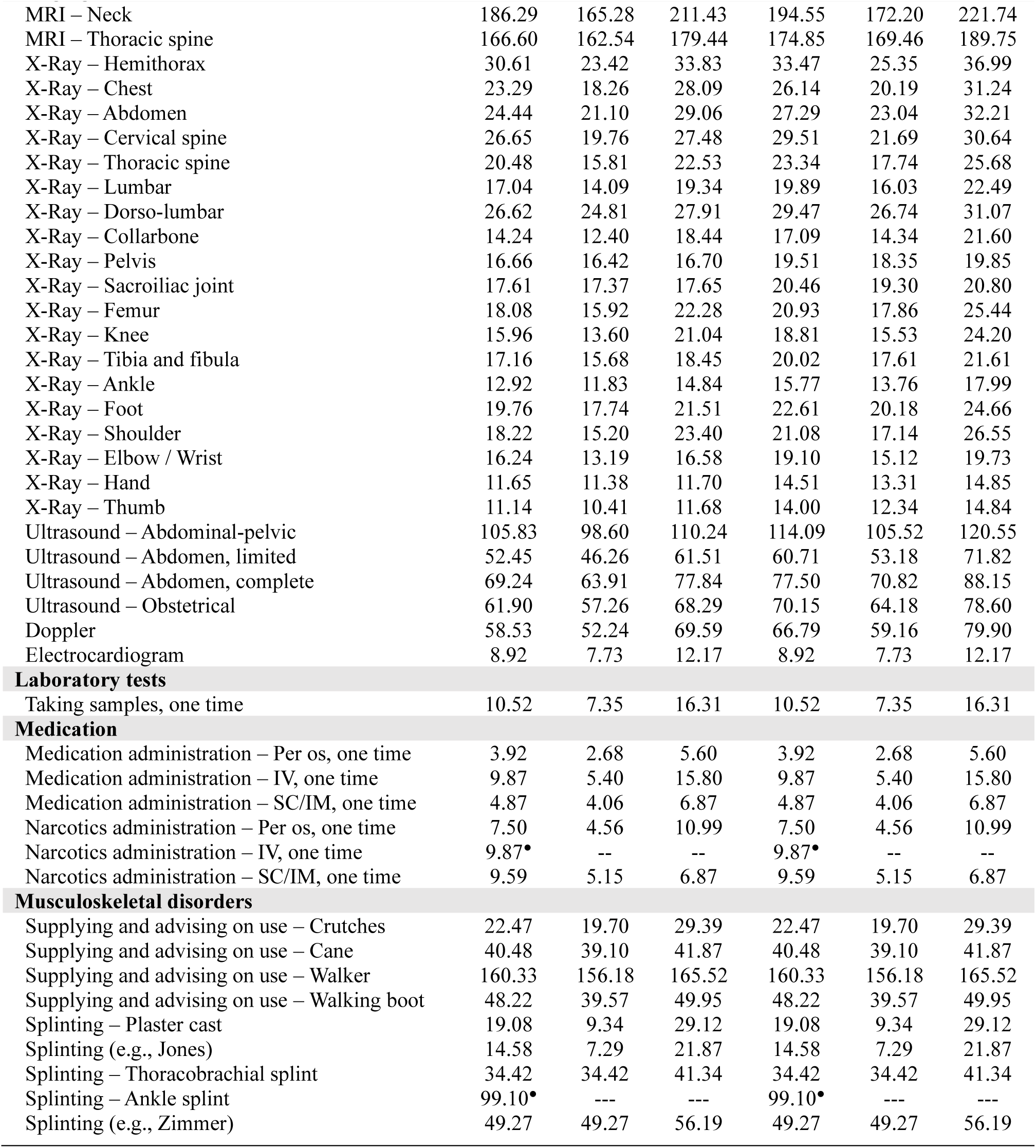
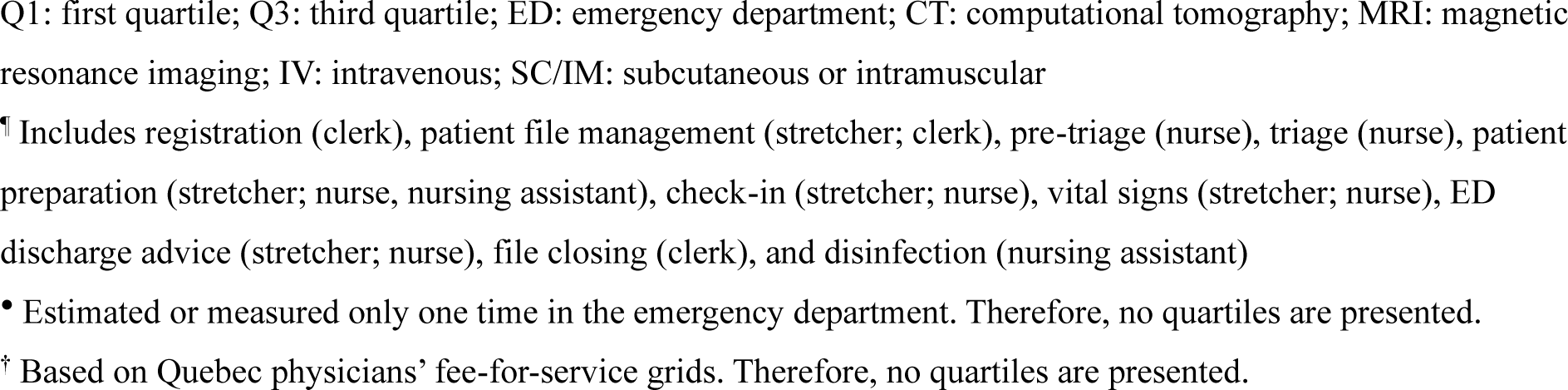
Unit cost of care processes used to calculate the cost of emergency department visits.

| Care process | Costs (in 2019 Canadian dollars) |  |  |  |  |  |
| --- | --- | --- | --- | --- | --- | --- |
|  | Ambulatory |  |  | Stretcher |  |  |
|  | Median | Q1 | Q3 | Median | Q1 | Q3 |
| **ED check-in** |  |  |  |  |  |  |
| ED arrival<sup>¶</sup> - On feet | 15.37 | 11.54 | 20.77 | 49.18 | 35.35 | 65.08 |
| ED arrival<sup>¶</sup> - Ambulance | 12.76 | 9.30 | 16.37 | 46.58 | 33.12 | 60.68 |
| ED arrival<sup>¶</sup> - With standing orders | 17.28 | 12.63 | 22.47 | 51.10 | 36.45 | 66.77 |
| Triage reassessment | 2.26 | 1.07 | 3.63 | 2.26 | 1.07 | 3.63 |
| ED physician initial assessment | 48.62 | 28.11 | 76.96 | 53.59 | 31.46 | 85.36 |
| ED physician reassessment | 38.16 | 23.24 | 58.92 | 44.86 | 33.34 | 60.04 |
| Physiotherapist initial assessment | 80.01<sup>•</sup> | -- | -- | 120.01<sup>•</sup> | -- | -- |
| Physiotherapist reassessment | -- | -- | -- | 20.00<sup>•</sup> | -- | -- |
| Consultant assessment – Internist | 215.90<sup>†</sup> | -- | -- | 215.90<sup>†</sup> | -- | -- |
| Consultant assessment – Social worker | --- | -- | -- | 83.40<sup>•</sup> | -- | -- |
| Consultant assessment – Occupational therapist | --- | -- | -- | 79.91<sup>•</sup> | -- | -- |
| Point-of-care ultrasound | 35.73 | 28.52 | 51.20 | 35.73 | 28.52 | 51.20 |
| **Imaging** |  |  |  |  |  |  |
| CT scan – Angiography | 137.89 | 127.16 | 151.15 | 146.14 | 134.08 | 161.46 |
| CT scan – Abdominal pelvic | 114.91 | 106.75 | 131.19 | 123.16 | 113.67 | 141.50 |
| CT scan – Pelvic | 87.91 | 79.75 | 104.19 | 96.16 | 86.67 | 114.50 |
| CT scan – Head | 58.84 | 53.43 | 70.97 | 67.10 | 60.35 | 81.29 |
| CT scan – Cervical spine | 87.44 | 81.31 | 98.76 | 95.69 | 88.23 | 109.07 |
| CT scan – Lumbar spine | 75.70 | 70.34 | 88.42 | 83.96 | 77.26 | 98.74 |
| CT scan – Ankle | 64.95 | 56.32 | 70.15 | 73.20 | 63.23 | 80.46 |
| CT scan – Foot | 70.13 | 62.22 | 84.63 | 78.38 | 69.14 | 94.94 |

**Table 2. Unit cost of care processes used to calculate the cost of emergency department visits (continued)**
| Care process | Costs (in 2019 Canadian dollars) |  |  |  |  |  |
| --- | --- | --- | --- | --- | --- | --- |
|  | Ambulatory |  |  | Stretcher |  |  |
|  | Median | Q1 | Q3 | Median | Q1 | Q3 |
| **Imaging (continued)** |  |  |  |  |  |  |
| MRI – Neck | 186.29 | 165.28 | 211.43 | 194.55 | 172.20 | 221.74 |
| MRI – Thoracic spine | 166.60 | 162.54 | 179.44 | 174.85 | 169.46 | 189.75 |
| X-Ray – Hemithorax | 30.61 | 23.42 | 33.83 | 33.47 | 25.35 | 36.99 |
| X-Ray – Chest | 23.29 | 18.26 | 28.09 | 26.14 | 20.19 | 31.24 |
| X-Ray – Abdomen | 24.44 | 21.10 | 29.06 | 27.29 | 23.04 | 32.21 |
| X-Ray – Cervical spine | 26.65 | 19.76 | 27.48 | 29.51 | 21.69 | 30.64 |
| X-Ray – Thoracic spine | 20.48 | 15.81 | 22.53 | 23.34 | 17.74 | 25.68 |
| X-Ray – Lumbar | 17.04 | 14.09 | 19.34 | 19.89 | 16.03 | 22.49 |
| X-Ray – Dorso-lumbar | 26.62 | 24.81 | 27.91 | 29.47 | 26.74 | 31.07 |
| X-Ray – Collarbone | 14.24 | 12.40 | 18.44 | 17.09 | 14.34 | 21.60 |
| X-Ray – Pelvis | 16.66 | 16.42 | 16.70 | 19.51 | 18.35 | 19.85 |
| X-Ray – Sacroiliac joint | 17.61 | 17.37 | 17.65 | 20.46 | 19.30 | 20.80 |
| X-Ray – Femur | 18.08 | 15.92 | 22.28 | 20.93 | 17.86 | 25.44 |
| X-Ray – Knee | 15.96 | 13.60 | 21.04 | 18.81 | 15.53 | 24.20 |
| X-Ray – Tibia and fibula | 17.16 | 15.68 | 18.45 | 20.02 | 17.61 | 21.61 |
| X-Ray – Ankle | 12.92 | 11.83 | 14.84 | 15.77 | 13.76 | 17.99 |
| X-Ray – Foot | 19.76 | 17.74 | 21.51 | 22.61 | 20.18 | 24.66 |
| X-Ray – Shoulder | 18.22 | 15.20 | 23.40 | 21.08 | 17.14 | 26.55 |
| X-Ray – Elbow / Wrist | 16.24 | 13.19 | 16.58 | 19.10 | 15.12 | 19.73 |
| X-Ray – Hand | 11.65 | 11.38 | 11.70 | 14.51 | 13.31 | 14.85 |
| X-Ray – Thumb | 11.14 | 10.41 | 11.68 | 14.00 | 12.34 | 14.84 |
| Ultrasound – Abdominal-pelvic | 105.83 | 98.60 | 110.24 | 114.09 | 105.52 | 120.55 |
| Ultrasound – Abdomen, limited | 52.45 | 46.26 | 61.51 | 60.71 | 53.18 | 71.82 |
| Ultrasound – Abdomen, complete | 69.24 | 63.91 | 77.84 | 77.50 | 70.82 | 88.15 |
| Ultrasound – Obstetrical | 61.90 | 57.26 | 68.29 | 70.15 | 64.18 | 78.60 |
| Doppler | 58.53 | 52.24 | 69.59 | 66.79 | 59.16 | 79.90 |
| Electrocardiogram | 8.92 | 7.73 | 12.17 | 8.92 | 7.73 | 12.17 |
| **Laboratory tests** |  |  |  |  |  |  |
| Taking samples, one time | 10.52 | 7.35 | 16.31 | 10.52 | 7.35 | 16.31 |
| **Medication** |  |  |  |  |  |  |
| Medication administration – Per os, one time | 3.92 | 2.68 | 5.60 | 3.92 | 2.68 | 5.60 |
| Medication administration – IV, one time | 9.87 | 5.40 | 15.80 | 9.87 | 5.40 | 15.80 |
| Medication administration – SC/IM, one time | 4.87 | 4.06 | 6.87 | 4.87 | 4.06 | 6.87 |
| Narcotics administration – Per os, one time | 7.50 | 4.56 | 10.99 | 7.50 | 4.56 | 10.99 |
| Narcotics administration – IV, one time | 9.87* | -- | -- | 9.87* | -- | -- |
| Narcotics administration – SC/IM, one time | 9.59 | 5.15 | 6.87 | 9.59 | 5.15 | 6.87 |
| **Musculoskeletal disorders** |  |  |  |  |  |  |
| Supplying and advising on use – Crutches | 22.47 | 19.70 | 29.39 | 22.47 | 19.70 | 29.39 |
| Supplying and advising on use – Cane | 40.48 | 39.10 | 41.87 | 40.48 | 39.10 | 41.87 |
| Supplying and advising on use – Walker | 160.33 | 156.18 | 165.52 | 160.33 | 156.18 | 165.52 |
| Supplying and advising on use – Walking boot | 48.22 | 39.57 | 49.95 | 48.22 | 39.57 | 49.95 |
| Splinting – Plaster cast | 19.08 | 9.34 | 29.12 | 19.08 | 9.34 | 29.12 |
| Splinting (e.g., Jones) | 14.58 | 7.29 | 21.87 | 14.58 | 7.29 | 21.87 |
| Splinting – Thoracobrachial splint | 34.42 | 34.42 | 41.34 | 34.42 | 34.42 | 41.34 |
| Splinting – Ankle splint | 99.10* | --- | --- | 99.10* | --- | --- |
| Splinting (e.g., Zimmer) | 49.27 | 49.27 | 56.19 | 49.27 | 49.27 | 56.19 |

Q1: first quartile; Q3: third quartile; ED: emergency department; CT: computational tomography; MRI: magnetic resonance imaging; IV: intravenous; SC/IM: subcutaneous or intramuscular
<sup>¶</sup> Includes registration (clerk), patient file management (stretcher; clerk), pre-triage (nurse), triage (nurse), patient preparation (stretcher; nurse, nursing assistant), check-in (stretcher; nurse), vital signs (stretcher; nurse), ED discharge advice (stretcher; nurse), file closing (clerk), and disinfection (nursing assistant)
• Estimated or measured only one time in the emergency department. Therefore, no quartiles are presented.
<sup>†</sup> Based on Quebec physicians' fee-for-service grids. Therefore, no quartiles are presented.

### Cost associated with the ED stay

Based on field measurements, it is estimated that only 40% of a nurse and nursing assistant’s hourly wages are captured by activities performed directly on patients in stretchers. Thus, the average ED stay cost is obtained by 1) dividing the remaining 60% of the nurse and nursing assistant’s hourly wage between the number of patients under their care (2.5 patient/hour/professional), and 2) multiplying the hourly patient cost obtained by the number of hours spent in the ED for each patient. This cost only applies to stretcher patients, since ambulatory patients do not require continuous monitoring by a nurse and/or nursing assistant.

Once the unit cost of each care process had been determined, the total cost of each patient’s ED visit was estimated by adding up the unit cost of each care process they required with the cost associated with their ED stay on a stretcher, if applicable.

### Data analyses and sensitivity analyses

#### Data analyses

All measured and estimated care process durations are presented as medians (Table S2). Quartiles are also presented for measured care process durations. The capacity cost rate of ED resources (Table 1) and unit costs of care processes (Table 2), as well as the average total cost of the ED visit per patient for each ED care model (Tables 4 and 5), are reported as mean costs. Confidence intervals (95%) for all costs presented were calculated using non-parametric Bootstrap methods (1,000 samples). Generalized linear models with a Gamma distribution and log links were used to determine whether there were significant differences between the average cost of an ED visit between the ED care models. Moreover, stratified analyses were performed to determine whether there was a significant difference in the average cost of an ED visit between the two care models studied according to sex, ED orientation (ambulatory or stretcher), and region of presenting MSKD. In line with economic evaluation guidelines, all absolute cost differences are reported, regardless of significance. All costs are reported in 2019 Canadian dollar values (CAD) (1 USD = 1.3363 CAD; March 29, 2019 – Bank of Canada). All statistical analyses were performed using SAS statistical analysis software (version 9.4, SAS Institute, Cary, North Carolina, USA).

#### Sensitivity analyses

Three different sensitivity analysis scenarios were carried out to represent the possible variability and uncertainty in costs associated with ED visits:

1. As the average ED visit costs were calculated using the median duration of care processes, the duration of each care process was varied in the first scenario (1st quartile, 3rd quartile).
2. In the second scenario, the duration of the physiotherapist’s assessment was varied until a cut-off assessment duration was reached where the average ED visit cost was the same between the two care models being compared (emergency physician vs. physiotherapist and emergency physician).
3. The third sensitivity analysis scenario was used to explore the average ED visit cost of a hypothetical ED care model where the physiotherapist could have autonomously managed patients when they deemed a consultation with the emergency physician was unnecessary. When necessary, the patient would have been referred to the emergency physician (e.g., prescription of medication and/or imaging tests).

## Results

### Participants’ sociodemographic characteristics

Sociodemographic characteristics of included participants are shown in Table 3:

**Table 3.** Participants’ sociodemographic characteristics (n=78) *.

| Characteristics | EP<br>n=38 | PT + EP<br>n=40 |
| --- | --- | --- |
| Number of participants, *n* (%) | 38 (48.7) | 40 (51.3) |
| Age (yr), mean (SD) | 44.1 (17.2) | 36.6 (17.3) |
| Sex, *n* females (%) | 12 (31.6) | 22 (55.0) |
| Triage category<sup>¶</sup> in ED, *n* (%) |  |  |
| Urgent (P3) | 16 (42.1) | 16 (40.0) |
| Semi urgent (P4) | 21 (55.3) | 24 (60.0) |
| Non urgent (P5) | 1 (2.6) | 0 (0.0) |
| ED length of stay (h), mean (SD) | 7.1 (4.9) | 6.7 (4.5) |
| Other health condition, yes (%) | 23 (60.5) | 26 (65.0) |

EP: emergency physician; PT: physiotherapist; yr: year; SD: standard deviation; ED: emergency department; h: hour
\* A version of this table containing more information on participants can be found in the supplemental document *Table S3*.
<sup>¶</sup> According to the Canadian Triage and Acuity Scale (CTAS)
More information on the participants included in this study can be found in the following article:
*Gagnon R, Perreault K, Berthelot S, et al. Direct-access physiotherapy to help manage patients with musculoskeletal disorders in an emergency department: Results of a randomized controlled trial. Acad Emerg Med 2021;28(8):848-858. DOI: 10.1111/acem.14237.*

**Table 4.** Average cost of an emergency department visit by care model.

| Care processes | Costs (in 2019 Canadian dollars) (n=78) |  |  |  | Sensitivity analysis - PT |  |
| --- | --- | --- | --- | --- | --- | --- |
|  | EP<br>n=38 | PT + EP<br>n=40 | Absolute difference<br>(95% CI) | p | Costs* – Median<br>time values | Absolute difference<br>(95% CI) |
| Average cost of the ED visit | 245.14<br>(169.46, 336.72) | 267.08<br>(212.75, 346.40) | 21.94<br>(-87.33, 132.63) | .60 | 194.38<br>(161.50, 234.34) | 50.76<br>(-156.91, 37.54) |
| **ED check-in<sup>¶</sup>** | **116.25**<br>**(96.59, 136.29)** | **192.34**<br>**(171.39, 223.03)** | **76.09**<br>**(43.90, 109.56)** | **<.01** | 136.31<br>(119.16, 153.43) | 20.06<br>(-9.39, 46.71) |
| ED stay<sup>•</sup> | 61.99<br>(22.56, 113.25) | 36.57<br>(8.63, 74.72) | 25.42<br>(-33.49, 85.36) | .99 | 24.50<br>(5.12, 50.66) | 37.49<br>(-14.36, 95.99) |
| Imaging | 37.06<br>(22.94, 53.67) | 16.88<br>(10.31, 26.11) | 20.18<br>(-37.99, -2.57) | .15 | 11.74<br>(8.18, 16.06) | 25.32<br>(-41.24, -10.47) |
| Laboratory tests | 6.53<br>(0.59, 17.51) | 5.16<br>(0.42, 13.96) | 1.37<br>(-13.84, 10.45) | >.99 | 4.57<br>(0.00, 13.05) | 1.96<br>(-13.84, 8.87) |
| Medication | 5.51<br>(2.57, 9.48) | 2.44<br>(0.95, 4.50) | 3.07<br>(-7.25, 0.59) | .90 | 2.44<br>(0.92, 4.18) | 3.07<br>(-7.11, 0.49) |
| Musculoskeletal disorders<sup>†</sup> | 17.80<br>(7.05, 30.28) | 13.68<br>(7.75, 20.00) | 4.12<br>(-17.52, 8.10) | .96 | 14.81<br>(8.97, 21.69) | 2.99<br>(-16.23, 9.64) |

CI: confidence interval; EP: emergency physician; PT: physiotherapist; ED: emergency department
\*Costs are presented as mean (95% confidence intervals obtained via non-parametric Bootstrap).
<sup>¶</sup> Includes ED arrival (clerk, nurse, nursing assistant), triage reassessment (if needed; nurse), assessment (emergency physician; if needed: physiotherapist, internist, social worker, occupational therapist), reassessment (if needed; emergency physician, physiotherapist), point-of-care ultrasound (if needed; emergency physician).
<sup>•</sup> Since all the parameters needed to calculate the ED stay cost are fixed (hours billed, patient ratio, ED length of stay), the average costs obtained are the same for all three scenarios (Median, Q1 and Q3 durations).
<sup>†</sup> Musculoskeletal disorders care processes include supplying and advising on use for walking aids and orthoses (crutches, cane, walker or walking boot), and splinting (plaster cast, Jones splint, thoracobrachial splint, ankle splint or Zimmer splint).

**Table 5.**
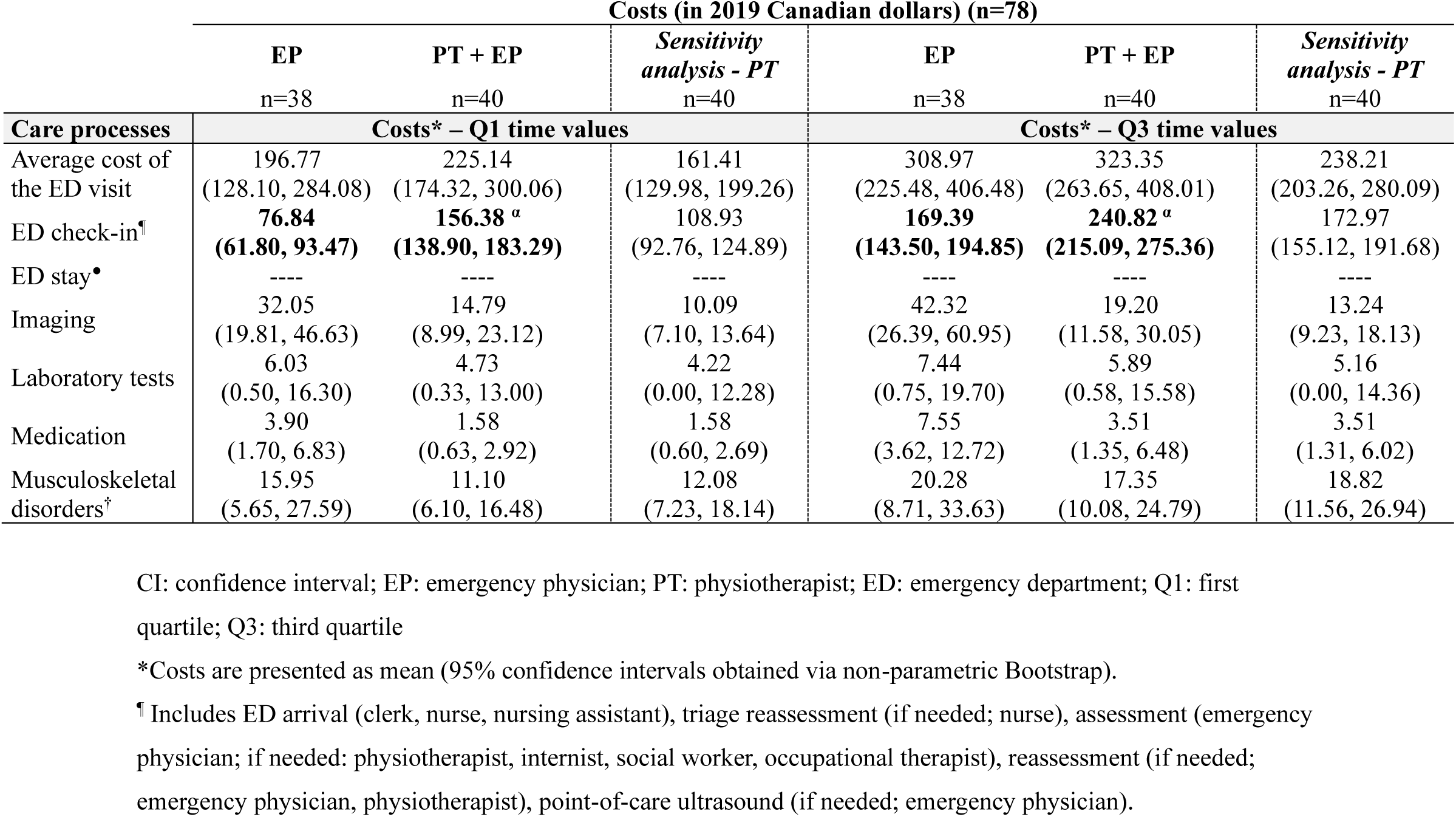
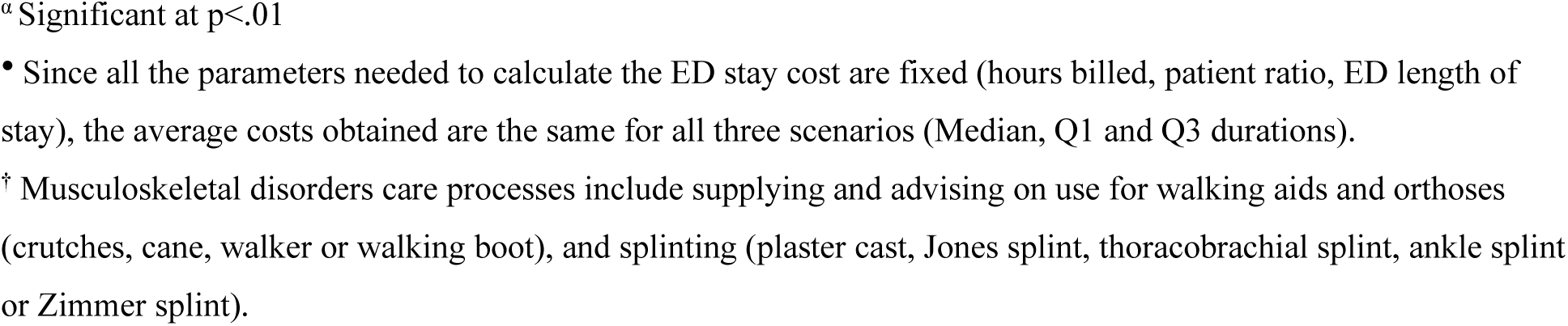
Average cost of an emergency department visit by care model according to different care process durations (sensitivity analyses)

| Care processes | Costs (in 2019 Canadian dollars) (n=78) |  |  |  |  |  |
| --- | --- | --- | --- | --- | --- | --- |
|  | EP<br>n=38 | PT + EP<br>n=40 | Sensitivity<br>analysis - PT<br>n=40 | EP<br>n=38 | PT + EP<br>n=40 | Sensitivity<br>analysis - PT<br>n=40 |
|  | Costs* – Q1 time values |  |  | Costs* – Q3 time values |  |  |
| Average cost of the ED visit | 196.77<br>(128.10, 284.08) | 225.14<br>(174.32, 300.06) | 161.41<br>(129.98, 199.26) | 308.97<br>(225.48, 406.48) | 323.35<br>(263.65, 408.01) | 238.21<br>(203.26, 280.09) |
| ED check-in<sup>¶</sup> | **76.84**<br>**(61.80, 93.47)** | **156.38<sup>a</sup>**<br>**(138.90, 183.29)** | 108.93<br>(92.76, 124.89) | **169.39**<br>**(143.50, 194.85)** | **240.82<sup>a</sup>**<br>**(215.09, 275.36)** | 172.97<br>(155.12, 191.68) |
| ED stay<sup>•</sup> | ---- | ---- | ---- | ---- | ---- | ---- |
| Imaging | 32.05<br>(19.81, 46.63) | 14.79<br>(8.99, 23.12) | 10.09<br>(7.10, 13.64) | 42.32<br>(26.39, 60.95) | 19.20<br>(11.58, 30.05) | 13.24<br>(9.23, 18.13) |
| Laboratory tests | 6.03<br>(0.50, 16.30) | 4.73<br>(0.33, 13.00) | 4.22<br>(0.00, 12.28) | 7.44<br>(0.75, 19.70) | 5.89<br>(0.58, 15.58) | 5.16<br>(0.00, 14.36) |
| Medication | 3.90<br>(1.70, 6.83) | 1.58<br>(0.63, 2.92) | 1.58<br>(0.60, 2.69) | 7.55<br>(3.62, 12.72) | 3.51<br>(1.35, 6.48) | 3.51<br>(1.31, 6.02) |
| Musculoskeletal disorders<sup>†</sup> | 15.95<br>(5.65, 27.59) | 11.10<br>(6.10, 16.48) | 12.08<br>(7.23, 18.14) | 20.28<br>(8.71, 33.63) | 17.35<br>(10.08, 24.79) | 18.82<br>(11.56, 26.94) |

CI: confidence interval; EP: emergency physician; PT: physiotherapist; ED: emergency department; Q1: first quartile; Q3: third quartile
\*Costs are presented as mean (95% confidence intervals obtained via non-parametric Bootstrap).
<sup>¶</sup> Includes ED arrival (clerk, nurse, nursing assistant), triage reassessment (if needed; nurse), assessment (emergency physician; if needed: physiotherapist, internist, social worker, occupational therapist), reassessment (if needed; emergency physician, physiotherapist), point-of-care ultrasound (if needed; emergency physician).
<sup>a</sup> Significant at $p < .01$
- Since all the parameters needed to calculate the ED stay cost are fixed (hours billed, patient ratio, ED length of stay), the average costs obtained are the same for all three scenarios (Median, Q1 and Q3 durations).
<sup>†</sup> Musculoskeletal disorders care processes include supplying and advising on use for walking aids and orthoses (crutches, cane, walker or walking boot), and splinting (plaster cast, Jones splint, thoracobrachial splint, ankle splint or Zimmer splint).

A more complete version of this table, in which participants are stratified according to their ED orientation (ambulatory or stretcher), can also be found in the supplemental document Table S3.

The distribution of the different triage categories, length of stay in the ED and the proportion of participants with at least one comorbidity were very similar between the two care models. However, the average age of the intervention group was lower, and included more women than the control group (Table 3).

### Average cost of an emergency department visit

The average costs per patient of an ED visit for each care model are presented in Table 4. A stratification of these average costs according to ED orientation (i.e., ambulatory or stretcher) can also be found in Tables S4 and S5.

No significant differences were observed between the two care models in terms of the average costs of the ED visit and for costs associated with the different types of care processes, except for the cost of ED check-in, where management by a primary contact physiotherapist and an emergency physician cost significantly more (+76.09 CAD/patient; 95CI: $43.90, $109.56; p<.01) than usual management by an emergency physician (Table 4). However, when stratified according to ED orientation, the average cost of the ED visit was significantly higher (+66.42 CAD/patient; 95CI: $32.86, $97.65; p<.01) for ambulatory participants in the intervention group (Table S4). Absolute differences were observed in the average costs associated with the ED stay (-25.42 CAD/patient; 95CI: $-33.49, $85.36), imaging tests (-20.18 CAD/patient; 95CI: $-37.99, $-2.57), laboratory tests (-1.37 CAD/patient; 95CI: $-13.84, $10.45), medication (-3.07 CAD/patient; 95CI: $-7.25, $0.59) and MSKDs (-4.12 CAD/patient; 95CI: $-17.52, $8.10) (Table 4). Observed absolute differences were generally in favor of the intervention group but did not reach statistical significance. Despite the observed absolute differences, the average cost of the ED visit was higher in the intervention group than in the control group (+21.94 CAD/patient; 95CI: $-87.33, $132.63) (Table 4). Stratified analyses for sex and type of MSKD (extremity/spine) showed similar results to the analyses of the whole study population (data not shown).

### Sensitivity analyses

#### Care processes duration

The average costs per type of care process and per care model obtained using first and third quartiles durations are shown in Table 5.

Once again, average costs of the ED visit and ED check-in were higher in the intervention group, while costs for imaging, laboratory tests, medication and MSKDs were in favor of the intervention group, but this trend was non-significant (Table 5).

#### Physiotherapist’s assessment duration

Based on the obtained values of average cost of an ED visit per care model (Table 4), the duration of the initial primary contact physiotherapist assessment would have to be reduced by 16.5 minutes to obtain an identical average cost between the two groups (First quartile scenario: 21.3 minutes, Third quartile scenario: 10.8 minutes).

#### Hypothetical care model (physiotherapist alone)

Table 4 also shows the average cost of an ED visit under the care model where the physiotherapist would have the capacity to independently manage certain patients (including ED discharge). Within this scenario, although not statistically significant, the average cost of an ED visit would be 50.76 CAD/patient (95CI: $-156.91, $37.54) lower in the group managed by a physiotherapist independently than in the group managed in the usual way by an emergency physician. Average costs per patient for all types of care processes used would be lower in this care model, except for ED check-in (+20.06 CAD/patient; 95CI: $-9.39, $46.71).

## Discussion

This study represents the first North American study to report average costs of managing persons presenting with a MSKD in the ED. Our results show that the average cost of an ED visit involving a primary contact physiotherapist and an emergency physician was equivalent to a visit under usual care (emergency physician alone). However, stratified analyses suggest that the combination of a primary contact physiotherapist and an emergency physician is more expensive than usual care for ambulatory ED patients. This can be explained by the fact that the intervention group was managed by two healthcare professionals, rather than just one, as in the control group, which significantly increased ED check-in costs.

Recent work by organizations such as the Australian Physiotherapy Association and World Physiotherapy describe physiotherapists who independently assess, diagnose, manage (which may include the performance of medical delegated acts), and discharge patients in contexts such as the ED as advanced practice physiotherapists.^33,34^ Nonetheless, aspects of what has been labelled as advanced practice may not be allowed within existing regulatory or legislative frameworks.^34^ Although the bylaws in effect at the hospital where the RCT was conducted did not allow it, in Canada, the physiotherapist is a healthcare professional who can independently manage patients presenting with a wide range of conditions, including minor MSKDs, and in a variety of clinical settings.^32^ Results of our sensitivity analyses show that the implementation of an ED care model with fully autonomous management by a physiotherapist would enable the Public Payer to pay an average ED check-in cost similar to that of the control group, in addition to saving more than 50 CAD/patient per ED visit. More and more studies report that involving different professionals in the ED improves patients’ outcomes,^35^ care quality^36^ and work conditions,^37^ increases patients’ satisfaction,^38^ and reduces clinical errors^38^ and ED length of stay.^37^ The findings of this study highlight that EDs could benefit from greater integration of various healthcare professionals, such as primary contact physiotherapists, at a cost similar to that of usual care by an emergency physician.

Although the average difference in costs of certain types of care processes (i.e., ED stay, imaging and laboratory tests, medication, and MSKDs) was generally lower when managed by a primary contact physiotherapist and an emergency physician, no significant difference was observed. These results may highlight a divergence from the scientific literature on the subject, which reports that the addition of a physiotherapist in the ED could lead to a reduction in time waited before receiving care, ED length of stay, as well as in the use of imaging tests and prescription medication during the ED visit.^18–20^ While it could be expected that the reduced use of ED resources and services reported in the literature would lead to a significant reduction in ED costs, this was not observed in our study. This may be explained in part by the fact that the data were derived from a pilot RCT with a small sample size, and by the high variability of the care trajectories included; it is therefore possible that some higher or lower ED visit costs may have caused the averages to vary significantly. It is also important to note that the remuneration received by physicians varies from one country to another, and therefore from one study to another. As this is one of the most important cost drivers in the ED, it is possible that the cost of the medical assessment may increase or decrease the average cost of the ED visit, depending on prevailing medical practices and remuneration.

To the best of our knowledge, only two other studies have however directly examined the costs associated with the management of minor MSKDs by a physiotherapist in the ED. The first, by McClellan et al^39^, concluded that autonomous physiotherapy management by an extended scope physiotherapist is at least as costly as usual management by an ED physician while the second, by Richardson et al^40^, concluded that autonomous management by a physiotherapist (i.e., initial physiotherapy assessment and management) resulted in costs equivalent to usual care. These findings are aligned with the results obtained in our analyses, where the care models resulted in a generally favorable, but not statistically different, cost difference when compared to usual care. However, some of the scenarios studied by McClellan et al. (e.g., costs limited to those incurred by the hospital system, median and not average costs) rather support an equivalent or even lower cost for the autonomous physiotherapist care model. Their study also has some notable limitations, in that the calculation of costs related to consultations with health professionals is based on non-verified consultation times, and certain types of MSKDs were excluded, including fractures, MSKDs that occurred more than 72 hours prior, and MSKDs requiring opioid analgesia. Our study addresses some of these limitations, in that it is based on verified times, and includes all types of minor MSKDs that can be managed in an ED. The second study, by Richardson et al. has similar limitations, as the authors also chose to exclude certain types of MSKDs (i.e., fractures and MSKDs requiring immediate analgesia), thus limiting the external validity of the included sample. These two studies were also carried out in a healthcare system where the legislative framework allows physiotherapists to practice in advanced practice roles, including acts outside of the usual scope of practice such as autonomous imaging requests and prescription of some types of medication.^41,42^ As increasing professional roles is one of the suggested solutions for reducing ED overcrowding,^8,43^ it would be interesting to evaluate the impact on management costs of certain legislative changes in the roles and responsibilities of primary contact physiotherapists.

## Limitations

There are a number of limitations to this study, which we feel are important to mention. Firstly, the costs reported in this manuscript show considerable variability. This heterogeneity could be due in part to the fact that, given the pragmatic nature of the RCT, the ED care pathways were not standardized. This lack of standardization may have had an upward or downward influence on the number of care processes used for the same disorder, and therefore on the average costs calculated. In addition, for the same MSKD, there was considerable variability in clinical presentations and severity levels within the recruited sample. This variability was partially considered in our application of the TDABC method via the first scenario of the sensitivity analyses, where we varied the duration of care processes. Although important, a sensitivity analysis of this type does not entirely reflect the variability present in a care trajectory on an individual basis.

However, this same heterogeneity also represents one of this study’s strengths, in that it increases the external validity of the costing method used, and thus facilitates its application in any clinical setting other than the ED managing persons presenting with minor MSKDs. In addition, the adaptation of the TDABC method to the realities of the ED and the management of MSKDs was carried out by healthcare professionals with a good knowledge of the condition of interest, in addition to rigorous training in health economics. Finally, two other significant strengths of this study are that it includes all costs incurred, as well as exhaustively detailing the adaptation of the TDABC method to the realities of managing MSKDs in the ED.

## Conclusions

This study is a first step towards a better understanding of the care processes used and the costs incurred by the Public Payer for the management of persons presenting with MSKDs in the ED. Our results suggest that the average cost of an ED visit for a MSKD under a care model consisting of management by a primary contact physiotherapist and an emergency physician is equivalent to the average cost of an ED visit under the usual care model (emergency physician only). Sensitivity analyses showed that a care model where the physiotherapist manages patients alone could potentially result in savings for the Public Payer. Although important, management costs are not the only effectiveness metrics that should be considered when planning new ED care models. Indeed, improving healthcare organization should go beyond costs and determine whether the innovative care models put in place enable effectiveness gains in areas that are significant for persons visiting the ED (e.g., pain and function levels, quality of life). Given the healthcare system’s limited resources, it will also be relevant in the future to measure the overall efficiency of these new ED care models, to ensure that the money invested by the Public Payer is spent on care models that are effective.

## Data Availability

All data produced in the present study are available upon reasonable request to the authors.

## Author contributions

RG, KP, JRG, LJH, and SB were involved in study concept and design. RG, KP, JRG, LJH, and SB were involved in acquisition of the data. RG, KP, JRG, LJH, and SB were involved in analysis and interpretation of the data. RG drafted the manuscript. KP, JRG, LJH, and SB were involved in critical revision of the manuscript for important intellectual content. JRG and SB provided statistical expertise. RG, KP, JRG, and LJH were involved in acquisition of funding.

## Prior Presentations

Part of this work was presented at the Canadian Association for Health Services and Policy Research (CAHSPR) annual conference (Ottawa, Canada; May 17, 2024) and the North American Primary Care Research Group (NAPCRG) annual conference (Quebec, Canada; November 21, 2024).

## Acknowledgements

The authors would like to thank the following persons for their contributions: project participants, Antony Barabé, PT, physiotherapist at the *Centre hospitalier de l’Université Laval* (CHUL), and the entire team of managers at the *Direction des services multidisciplinaires* and *Direction des soins critiques* of the CHU de Québec – Université Laval (Marie-Christine Laroche, Catherine Van Neste, Jackie Chouinard, Marie-Claude Brodeur, Stéphane Tremblay) for their support throughout the implementation of the project and its realization.

## Supplemental Information linked to this paper

• Table S1

• Table S2

• Methods Supplement

• Table S3

• Table S4

• Table S5

