## Supplementary material for "Assessing innovative care models for musculoskeletal disorders’ management in the emergency department using Time-Driven Activity-Based Costing": Methods Supplement

**Methods Supplement** Method used to calculate the capacity cost rate for overhead

1. **Budget cost centers included in the calculation of overhead**
   a. Executive management
   b. Financial administration
   c. Personnel administration
   d. Professional services administration
   e. Supply and services
   f. Technical services administration
   g. Information technology
   h. Travel by ambulance to a palliative care home or for patients aged 65 and over
   i. Archives
   j. Telecommunications
   k. Food
   l. Laundry
   m. Hygiene and sanitation – Operational tasks
   n. Hygiene and sanitation – Functional tasks
   o. Biomedical waste management
   p. Hospital operations
   q. Security
   r. Maintenance of buildings and furniture / general equipment
2. **Example of overhead calculation**

Overhead – Emergency department

All calculations are based on data from the 2019 fiscal year.

CHU: CHU de Québec – Université Laval (5 academic hospitals located in Quebec City, Canada)

CHUL: *Centre hospitalier de l’Université Laval* (participating ED)

ED: Emergency department

$$Proportion of total CHU budget attributable to the ED=$$

$$\frac{5 EDs budget}{CHU's total budget}$$

$$CHU^{'}s overhead attributable to the ED=$$

$$CHU^{'}s total overhead* proportion of total CHU budget attributable to the ED$$

$$Proportion of patients visiting an ED at the CHU and seen at the study setting=$$

$$\frac{Number of patients seen at the CHUL}{Number of patients seen in the 5 CHU EDs}$$

$$Overhead specific to the CHUL's ED=$$

$$CHU^{'}s overhead attributable to the ED*$$

$$proportion of patients visiting the CHUL among all patients seen in the 5 EDs of the CHU$$
