## Supplemental Table 1 for "Assessing innovative care models for musculoskeletal disorders’ management in the emergency department using Time-Driven Activity-Based Costing"

**Table S1** Description of the steps and resources included in each care process

| **Care process** | **Included steps and resources** | |
| --- | --- | --- |
|  | **Ambulatory** | **Stretcher** |
| **ED check-in** |  |  |
| **ED arrival - On feet** | Registration (clerk)  Pre-triage (nurse)  Triage – arrival on feet (nurse)  Patient’s file closing (clerk)  ED room disinfection (nursing assistant) | Registration (clerk)  Patient’s file management – stretcher (clerk)  Pre-triage (nurse)  Triage – arrival on feet (nurse)  Preparation – stretcher patient (nurse)  Preparation – stretcher patient (nursing assistant)  Check-in – stretcher patient (nurse)  Vital signs (nurse)  ED discharge advice (nurse)  Patient’s file closing (clerk)  Disinfection – stretcher (nursing assistant) |
| **ED arrival - Ambulance** | Registration (clerk)  Triage – ambulance (nurse)  Patient’s file closing (clerk)  ED room disinfection (nursing assistant) | Registration (clerk)  Patient’s file management – stretcher (clerk) Triage – ambulance (nurse)  Preparation – stretcher patient (nurse)  Preparation – stretcher patient (nursing assistant)  Check-in – stretcher patient (nurse)  Vital signs (nurse)  ED discharge advice (nurse)  Patient’s file closing (clerk)  Disinfection – stretcher (nursing assistant) |
| **ED arrival - With triage**  **standing orders** | Registration (clerk)  Pre-triage (nurse)  Triage – With standing orders (nurse)  Patient’s file closing (clerk)  ED room disinfection (nursing assistant) | Registration (clerk)  Patient’s file management – stretcher (clerk)  Pre-triage (nurse)  Triage – With standing orders (nurse)  Preparation – stretcher patient (nurse)  Preparation – stretcher patient (nursing assistant)  Check-in – stretcher patient (nurse)  Vital signs (nurse)  ED departure advice (nurse)  Patient’s file closing (clerk)  Disinfection – stretcher (nursing assistant) |
| **Triage reassessment** | Triage reassessment (nurse) | Triage reassessment (nurse) |
| **ED physician initial**  **assessment** | Assessment (physician)  Clinical note (physician) | Assessment (physician)  Clinical note (physician) |
| **ED physician reassessment** | Reassessment (physician)  Clinical note (physician) | Reassessment (physician)  Clinical note (physician) |
| **Physiotherapist initial**  **assessment** | Assessment (physiotherapist)  Clinical note (physiotherapist) | Assessment (physiotherapist)  Clinical note (physiotherapist) |
| **Physiotherapist reassessment** | Reassessment (physiotherapist)  Clinical note (physiotherapist) | Reassessment (physiotherapist)  Clinical note (physiotherapist) |
| **Consultant assessment –**  **Internist** | Assessment (internist)  Clinical note (internist) | Assessment (internist)  Clinical note (internist) |

**Table S1** Description of the steps and resources included in each care process (continued)

| **Care process** | **Included steps and resources** | |
| --- | --- | --- |
|  | **Ambulatory** | **Stretcher** |
| **ED check-in (continued)** |  |  |
| **Consultant assessment –**  **Social worker** | Assessment (social worker)  Clinical note (social worker) | Assessment (social worker)  Clinical note (social worker) |
| **Consultant assessment –**  **Occupational therapist** | Assessment (occupational therapist)  Clinical note (occupational therapist) | Assessment (occupational therapist)  Clinical note (occupational therapist) |
| **Point-of-care ultrasound** | Objective assessment (physician)  Point-of-care ultrasound (physician) | Objective assessment (physician)  Point-of-care ultrasound (physician) |
| **Imaging** |  |  |
| **CT scan – Angiography** | Request – CT scan (clerk)  Request – hospital porter/nursing assistant (clerk)  Transport to imaging room and back – 2^nd^ floor (nursing assistant)  CT scan preparation – patient and device (imaging technician)  Exam (imaging technician and wear and tear of equipment)  Reinstallation – patient and device (imaging technician)  Interpretation (radiologist)  Results verification – imaging (clerk) | Request – CT scan (clerk)  Request – hospital porter/nursing assistant (clerk)  Transport to imaging room and back – 2^nd^ floor (nursing assistant)  CT scan preparation – patient and device (imaging technician)  Exam (imaging technician and wear and tear of equipment)  Reinstallation – patient and device (imaging technician)  Interpretation (radiologist)  Results verification – imaging (clerk) |
| **CT scan – Abdominal pelvic** |  |  |
| **CT scan – Pelvic** |  |  |
| **CT scan – Head** |  |  |
| **CT scan – Cervical spine** |  |  |
| **CT scan – Lumbar spine** |  |  |
| **CT scan – Ankle** |  |  |
| **CT scan – Foot** |  |  |
| **MRI – Neck** | Request – MRI (clerk)  Request – hospital porter/nursing assistant (clerk)  Transport to imaging room and back – 2^nd^ floor (nursing assistant)  MRI preparation – patient and device (imaging technician)  Exam (imaging technician and wear and tear of equipment)  Reinstallation – patient and device (imaging technician)  Interpretation (radiologist)  Results verification – imaging (clerk) | Request – MRI (clerk)  Request – hospital porter/nursing assistant (clerk)  Transport to imaging room and back – 2^nd^ floor (nursing assistant)  MRI preparation – patient and device (imaging technician)  Exam (imaging technician and wear and tear of equipment)  Reinstallation – patient and device (imaging technician)  Interpretation (radiologist)  Results verification – imaging (clerk) |
| **MRI – Thoracic spine** |  |  |
| **MRI – Lumbar spine** |  |  |

**Table S1** Description of the steps and resources included in each care process (continued)

| **Care process** | **Included steps and resources** | |
| --- | --- | --- |
|  | **Ambulatory** | **Stretcher** |
| **Imaging (continued)** |  |  |
| **X-Ray – Hemithorax** | Request – X-Ray (clerk)  Exam (imaging technician and wear and tear of equipment)  Interpretation (radiologist)  Results verification – imaging (clerk) | Request – X-Ray (clerk)  Request – hospital porter/nursing assistant (clerk)  Transport to imaging room and back – ED (nursing assistant)  Exam (imaging technician and wear and tear of equipment)  Interpretation (radiologist)  Results verification – imaging (clerk) |
| **X-Ray – Chest** |  |  |
| **X-Ray – Abdomen** |  |  |
| **X-Ray – Cervical spine** |  |  |
| **X-Ray – Thoracic spine** |  |  |
| **X-Ray – Lumbar** |  |  |
| **X-Ray – Dorso-lumbar** |  |  |
| **X-Ray – Collarbone** |  |  |
| **X-Ray – Pelvis** |  |  |
| **X-Ray – Sacroiliac joint** |  |  |
| **X-Ray – Femur** |  |  |
| **X-Ray – Knee** |  |  |
| **X-Ray – Tibia and fibula** |  |  |
| **X-Ray – Ankle** |  |  |
| **X-Ray – Foot** |  |  |
| **X-Ray – Shoulder** |  |  |
| **X-Ray – Elbow / Wrist** |  |  |
| **X-Ray – Hand** |  |  |
| **X-Ray – Thumb** |  |  |
| **Ultrasound – Abdominal-**  **pelvic** | Request – ultrasound (clerk)  Request – hospital porter/nursing assistant (clerk)  Transport to imaging room and back – 2^nd^ floor (nursing assistant)  Ultrasound preparation – patient and device (imaging technician)  Exam (imaging technician and wear and tear of equipment)  Reinstallation – patient and device (imaging technician)  Ultrasound verification (imaging technician and wear and tear of equipment)  Interpretation (radiologist)  Results verification – imaging (clerk) | Request – ultrasound (clerk)  Request – hospital porter/nursing assistant (clerk)  Transport to imaging room and back – 2^nd^ floor (nursing assistant)  Ultrasound preparation – patient and device (imaging technician)  Exam (imaging technician and wear and tear of equipment)  Reinstallation – patient and device (imaging technician)  Ultrasound verification (imaging technician and wear and tear of equipment)  Interpretation (radiologist)  Results verification – imaging (clerk) |
| **Ultrasound – Abdomen,**  **limited** |  |  |
| **Ultrasound – Abdomen,**  **complete** |  |  |
| **Ultrasound – Obstetrical** |  |  |

**Table S1** Description of the steps and resources included in each care process (continued)

| **Care process** | **Included steps and resources** | |
| --- | --- | --- |
|  | **Ambulatory** | **Stretcher** |
| **Imaging (continued)** |  |  |
| **Doppler** | Doppler request and planification (clerk)  Request – hospital porter/nursing assistant (clerk)  Transport to imaging room and back – 2^nd^ floor (nursing assistant)  Doppler preparation – patient and device (imaging technician)  Exam (imaging technician and wear and tear of equipment)  Reinstallation – patient and device (imaging technician)  Doppler verification (imaging technician and wear and tear of equipment)  Interpretation (radiologist)  Results verification – imaging (clerk) | Doppler request and planification (clerk)  Request – hospital porter/nursing assistant (clerk)  Transport to imaging room and back – 2^nd^ floor (nursing assistant)  Doppler preparation – patient and device (imaging technician)  Exam (imaging technician and wear and tear of equipment)  Reinstallation – patient and device (imaging technician)  Doppler verification (imaging technician and wear and tear of equipment)  Interpretation (radiologist)  Results verification – imaging (clerk) |
| **Electrocardiogram** | Request – electrocardiogram (clerk)  Exam (nursing assistant)  File delivery to the ambulatory section of the ED – electrocardiogram (nurse)  Interpretation (cardiologist)  Results verification – imaging (clerk) | Request – electrocardiogram (clerk)  Exam (nursing assistant)  File delivery to the ambulatory section of the ED – electrocardiogram (nurse)  Interpretation (cardiologist)  Results verification – imaging (clerk) |
| **Laboratory tests** |  |  |
| **Drawing laboratory tests,**  **one time** | Laboratory tests labelling (clerk)  Drawing tests (nurse)  Laboratory tests delivery (nursing assistant)  Results verification – Samples (clerk) | Laboratory tests labelling (clerk)  Drawing tests (nurse)  Laboratory tests delivery (nursing assistant)  Results verification – Samples (clerk) |
| **Medication** |  |  |
| **Medication administration –**  **Per os, one time** | Medication administration – Per os (nurse) | Medication administration – Per os (nurse) |
| **Medication administration –**  **IV, one time** | Medication administration – IV (nurse) | Medication administration – IV (nurse) |
| **Medication administration –**  **SC/IM, one time** | Medication administration – SC/IM (nurse) | Medication administration – SC/IM (nurse) |
| **Narcotics administration –**  **Per os, one time** | Narcotics administration – Per os (nurse)  Vital signs (nurse) | Narcotics administration – Per os (nurse)  Vital signs (nurse) |
| **Narcotics administration –**  **IV, one time** | Narcotics administration – IV (nurse)  Vital signs (nurse) | Narcotics administration – IV (nurse)  Vital signs (nurse) |
| **Narcotics administration –**  **SC/IM, one time** | Narcotics administration – SC/IM (nurse)  Vital signs (nurse) | Narcotics administration – SC/IM (nurse)  Vital signs (nurse) |

**Table S1** Description of the steps and resources included in each care process (continued)

| **Care process** | **Included steps and resources** | |
| --- | --- | --- |
|  | **Ambulatory** | **Stretcher** |
| **Musculoskeletal disorders** |  |  |
| **Supplying and advising on use**  **– Crutches** | Cost of walking aid (traceable supplies)  Education – Use of crutches (nurse) | Cost of walking aid (traceable supplies)  Education – Use of crutches (nurse) |
| **Supplying and advising on use**  **– Cane** | Cost of walking aid (traceable supplies)  Education – Use of cane (nurse) | Cost of walking aid (traceable supplies)  Education – Use of cane (nurse) |
| **Supplying and advising on use**  **– Walker** | Cost of walking aid (traceable supplies)  Education – Use of walker (nurse) | Cost of walking aid (traceable supplies)  Education – Use of walker (nurse) |
| **Supplying and advising on use**  **– Walking boot** | Cost of boot (traceable supplies)  Education – Walking boot (nurse) | Cost of boot (traceable supplies)  Education – Walking boot (nurse) |
| **Splinting – Plaster cast** | Cost is derived using the following equation:  [time taken splinting (nurse) * capacity cost rate (nurse)]  +  [time taken splinting (nurse) * capacity cost rate (overhead)]  +  [time taken splinting (nurse) * capacity cost rate (consumables)] | Cost is derived using the following equation:  [time taken splinting (nurse) * capacity cost rate (nurse)]  +  [time taken splinting (nurse) * capacity cost rate (overhead)]  +  [time taken splinting (nurse) * capacity cost rate (consumables)] |
| **Splinting (e.g., Jones)** | Cost is derived using the following equation:  [time taken splinting (nurse) * capacity cost rate (nurse)]  +  [time taken splinting (nurse) * capacity cost rate (overhead)]  +  [time taken splinting (nurse) * capacity cost rate (consumables)] | Cost is derived using the following equation:  [time taken splinting (nurse) * capacity cost rate (nurse)]  +  [time taken splinting (nurse) * capacity cost rate (overhead)]  +  [time taken splinting (nurse) * capacity cost rate (consumables)] |
| **Splinting – Thoracobrachial**  **splint** | Cost of splint (traceable supplies)  Splinting – Thoracobrachial splint (nurse) | Cost of splint (traceable supplies)  Splinting – Thoracobrachial splint (nurse) |
| **Splinting – Ankle splint** | Cost of splint (traceable supplies)  Splinting – Ankle splint (nurse) | Cost of splint (traceable supplies)  Splinting – Ankle splint (nurse) |
| **Splinting (e.g., Zimmer)** | Cost of splint (traceable supplies)  Splinting – Zimmer splint (nurse) | Cost of splint (traceable supplies)  Splinting – Zimmer splint (nurse) |

ED: emergency department; CT: computed tomography; MRI: magnetic resonance imaging; w/o: without; 2^nd^: second; IV: intravenous; SC/IM: subcutaneous or intramuscular
