## Supplemental Table 2 for "Assessing innovative care models for musculoskeletal disorders’ management in the emergency department using Time-Driven Activity-Based Costing"

**Table S2** Duration of the main care processes in the emergency department

| **Care process** | **Measured in the ED** | **Estimated by healthcare professionals** |
| --- | --- | --- |
|  | Mins, Median (Q1; Q3) | Mins, Median |
| **Clerk** |  |  |
| Registration | 2.3 (1.6; 2.8) |  |
| Patient’s file management – Stretcher |  | 2.0 |
| Results verification – Laboratory tests/Imaging | 0.4 (0.1; 0.4) |  |
| Request – Hospital porter | 0.7 (0.3; 0.8) |  |
| Request – Consultant |  | 1.0 |
| Request – Radiography | 0.6 (---; ---) |  |
| Request – Ultrasound/CT scan | 0.6 (---; ---) |  |
| Request – Electrocardiogram | 0.6 (0.4; 1.2) |  |
| Laboratory tests labelling | 0.9 (0.7; 2.4) |  |
| Patients’ file closing | 1.1 (0.7; 1.6) |  |
| Doppler request and planification | 4.8 (2.7; 4.9) |  |
| **Nurse** |  |  |
| Pre-triage | 1.0 (0.8; 1.3) |  |
| Triage – Arrival on feet | 7.1 (5.5; 9.3) |  |
| Triage – Ambulance | 6.2 (4.6; 7.4) |  |
| Triage – With standing orders | 8.5 (6.3; 10.5) |  |
| Vital signs | 5.1 (4.3; 5.9) |  |
| File delivery – Electrocardiogram | 1.4 (1.3; 2.3) |  |
| Triage reassessment | 1.6 (0.8; 2.6) |  |
| Preparation – Stretcher patient | 1.2 (0.8; 2.6) |  |
| Check-in – Stretcher patient | 8.7 (5.8; 11.7) |  |
| Monitoring – Stretcher patient | 36.0 (---; ---) / hour |  |
| ED departure advice | 2.2 (1.0; 3.6) |  |
| Medication administration – Per os | 2.8 (1.9; 4.1) |  |
| Medication administration –  Subcutaneous or intramuscular | 3.5 (2.9; 5.0) |  |
| Medication administration – IV | 7.1 (3.9; 11.4) |  |
| Narcotics administration – Per os | 5.4 (3.3; 8.0) |  |
| Narcotics administration – Subcutaneous | 6.9 (3.7; 10.0) |  |
| Narcotics administration – IV | 7.1 (---; ---) |  |
| Laboratory tests | 6.1 (4.1; 9.3) |  |
| Education – Use of crutches |  | 5.0 |
| Education – Use of cane |  | 4.0 |
| Education – Use of walker |  | 7.5 |
| Education – Walking boot |  | 10.0 |
| Splinting – Plaster cast | 13.1 (6.4; 20.0) |  |
| Splinting (e.g., Jones) |  | 10.0 |
| Splinting – Thoracobrachial splint |  | 5.0 |
| Splinting – Ankle splint |  | 3.0 |
| Splinting (e.g., Zimmer) |  | 5.0 |

**Table S2** Duration of the main care processes in the emergency department (continued)

| **Care process** | **Measured in the ED** | **Estimated by healthcare professionals** |
| --- | --- | --- |
|  | Mins, Median (Q1; Q3) | Mins, Median |
| **Nursing assistant** |  |  |
| Disinfection – ED room | 1.3 (0.9; 2.3) |  |
| Disinfection – Stretcher | 5.5 (4.2; 7.3) |  |
| Laboratory tests delivery |  | 1.0 |
| Preparation – Stretcher patient | 4.4 (2.6; 5.0) |  |
| Preparation – Transport to imaging | 1.2 (1.0; 1.7) |  |
| Transport to imaging room – ED | 1.1 (0.9; 1.3) |  |
| Transport to imaging room – Radiology on  second floor | 4.0 (3.5; 5.0) |  |
| **Physiotherapist** |  |  |
| Initial assessment – Ambulatory |  | 60.0 |
| Initial assessment – Stretcher |  | 90.0 |
| Reassessment – Stretcher |  | 15.0 |
| **Occupational Therapist** |  |  |
| Initial assessment – Stretcher |  | 90.0 |
| **Social Worker** |  |  |
| Initial assessment – Stretcher |  | 90.0 |
| **Emergency Physician** |  |  |
| Initial assessment – Ambulatory | 8.0 (4.6; 12.6) |  |
| Initial assessment – Stretcher | 8.8 (5.2; 14.0) |  |
| Reassessment – Ambulatory | 6.3 (3.8; 9.7) |  |
| Reassessment – Stretcher | 7.4 (5.5; 9.9) |  |
| Point-of-care ultrasound | 5.9 (4.7; 8.4) |  |
| Sampling – Synovial fluid |  | 15.0 |
| **CT scan** |  |  |
| Preparation – Patient and imaging device | 3.4 (2.5; 5.5) |  |
| Reinstallation – Patient and imaging device | 2.3 (1.7; 3.0) |  |
| Angiography | 4.4 (4.3; 5.7) |  |
| Abdominal pelvic | 4.6 (3.5; 6.9) |  |
| Pelvic | 4.6 (3.5; 6.9) |  |
| Head | 2.2 (2.0; 3.2) |  |
| Cervical spine | 4.5 (4.0; 5.2) |  |
| Lumbar spine | 2.7 (2.5; 3.9) |  |
| Ankle |  | 4.3 |
| Foot |  | 6.0 |
| **MRI** |  |  |
| Preparation – Patient and imaging device | 5.4 (4.5; 8.1) |  |
| Reinstallation – Patient and imaging device | 1.7 (1.6; 3.9) |  |
| Neck | 25.4 (17.1; 31.8) |  |
| Thoracic spine | 20.0 (19.3; 20.9) |  |

**Table S2** Duration of the main care processes in the emergency department (continued)

| **Care process** | **Measured in the ED** | **Estimated by healthcare professionals** |
| --- | --- | --- |
|  | Mins, Median (Q1; Q3) | Mins, Median |
| **X-Ray** |  |  |
| Hemithorax | 9.3 (6.4; 10.6) |  |
| Chest | 5.1 (3.1; 7.1) |  |
| Abdomen | 6.1 (4.8; 8.0) |  |
| Cervical spine | 11.5 (6.3; 12.2) |  |
| Thoracic spine | 8.6 (5.1; 10.1) |  |
| Lumbar | 5.9 (3.7; 7.6) |  |
| Dorso-lumbar | 12.6 (11.4; 13.6) |  |
| Clavicle |  | 5.0 |
| Pelvis / Sacroiliac joint |  | 4.5 |
| Femur |  | 8.0 |
| Knee | 6.3 (4.7; 10.3) |  |
| Tibia and fibula | 7.3 (6.3; 8.3) |  |
| Ankle | 4.0 (3.3; 5.4) |  |
| Foot | 5.3 (4.3; 6.6) |  |
| Shoulder | 6.8 (4.6; 10.8) |  |
| Elbow / Wrist | 6.6 (4.4; 6.8) |  |
| Hand | 3.0 (3.0; 3.0) |  |
| Thumb | 3.8 (3.4; 4.2) |  |
| **Ultrasound** |  |  |
| Preparation – Patient and imaging device | 1.7 (1.3; 2.6) |  |
| Reinstallation – Patient and imaging device | 2.2 (1.5; 2.6) |  |
| Verification | 5.3 (3.9; 7.8) |  |
| Abdominal-pelvic | 19.1 (15.5; 19.1) |  |
| Abdomen, limited | 8.1 (5.4; 12.2) |  |
| Abdomen, complete | 16.4 (14.4; 20.1) |  |
| Obstetrical | 13.3 (12.0; 15.1) |  |
| Doppler | 10.1 (9.0; 15.9) |  |
| **Other tests** |  |  |
| Electrocardiogram | 4.6 (3.8; 6.1) |  |

ED: emergency department; Mins: minutes; Q1: first quartile; Q3: third quartile; CT: computed tomography; IV: intravenous; 2^nd^: second; MRI: magnetic resonance imaging

More details about the method used to measure and estimate the times in this table, and some of the values in it, can be found in the following article by Berthelot et al:

*Berthelot S, Mallet M, Blais S, et al. Adaptation of time-driven activity-based costing to the evaluation of the efficiency of ambulatory care provided in the emergency department. J Am Coll Emerg Physicians Open 2022;3(4):e12778. DOI: 10.1002/emp2.12778.*
