## Supplemental Table 3 for "Assessing innovative care models for musculoskeletal disorders’ management in the emergency department using Time-Driven Activity-Based Costing"

**Table S3** Participants’ sociodemographic characteristics (n=78)

| **Characteristics** | **Ambulatory** | | **On stretcher** | |
| --- | --- | --- | --- | --- |
|  | **EP** | **PT + EP** | **EP** | **PT + EP** |
| Number of participants, *n* (%) | 28 (35.9) | 34 (43.6) | 10 (12.8) | 6 (7.7) |
| Age (yr), mean (SD) | 39.2 (16.0) | 35.0 (16.8) | 58.0 (12.6) | 45.3 (18.8) |
| Sex, *n* females (%) | 8 (28.6) | 18 (52.9) | 4 (40.0) | 4 (66.7) |
| Triage category^a^ in ED, *n* (%) |  |  |  |  |
| Urgent (P3) | 8 (28.6) | 10 (29.4) | 8 (80.0) | 6 (100.0) |
| Semi urgent (P4) | 19 (67.9) | 24 (70.6) | 2 (20.0) | 0 (0.0) |
| Non urgent (P5) | 1 (3.6) | 0 (0.0) | 0 (0.0) | 0 (0.0) |
| ED length of stay (h), mean (SD) | 6.4 (2.9) | 6.2 (3.9) | 9.2 (8.3) | 9.5 (7.0) |
| Other health condition, yes (%) | 13 (46.4) | 21 (61.8) | 10 (100.0) | 5 (83.3) |

EP: emergency physician; PT: physiotherapist; yr: year; SD: standard deviation; ED: emergency department; h: hour

^a^ According to the Canadian Triage and Acuity Scale (CTAS)

More information on the participants included in this study can be found in the following article:

*Gagnon R, Perreault K, Berthelot S, et al. Direct-access physiotherapy to help manage patients with musculoskeletal disorders in an emergency department: Results of a randomized controlled trial. Acad Emerg Med 2021;28(8):848-858. DOI: 10.1111/acem.14237.*
