## Supplemental Table 5 for "Assessing innovative care models for musculoskeletal disorders’ management in the emergency department using Time-Driven Activity-Based Costing"

**Table S5** Average cost of an emergency department visit by care model and orientation according to different care process durations (sensitivity analyses)

|  | **Costs (in 2019 Canadian dollars) (n=78)** | | | | | |
| --- | --- | --- | --- | --- | --- | --- |
|  | **EP** | **PT + EP** |  | **EP** | **PT + EP** |  |
| **Care processes** | n=28 | n=34 |  | n=28 | n=34 |  |
| **Ambulatory (n=62)** | **Costs^a^ – Q1 time values** | | ***p*** | **Costs^a^ – Q3 time values** | | ***p*** |
| **Average cost of the ED visit** | **96.12**  **(77.21, 114.87)** | **162.65**  **(147.68, 178.44)** | **<.01** | **189.17**  **(156.60, 224.00)** | **254.54**  **(226.98, 282.47)** | **.01** |
| **ED check-in**^b^ | **54.52**  **(46.36, 63.63)** | **134.15**  **(127.86, 139.86)** | **<.01** | **133.57**  **(113.06, 155.60)** | **213.86**  **(197.46, 228.45)** | **<.01** |
| ED stay^c^ | 0.00  (**---**) | 0.00  (**---**) | **---** | 0.00  (**---**) | 0.00  (**---**) | **---** |
| Imaging | 22.62  (13.79, 32.58) | 12.10  (8.80, 15.48) | .29 | 30.08  (18.71, 43.30) | 15.62  (11.25, 20.30) | .27 |
| Laboratory tests | 2.58  (0.00, 5.91) | 3.93  (0.00, 12.16) | .99 | 3.54  (0.00, 7.95) | 4.51  (0.00, 13.95) | .99 |
| Medication | 0.66  (0.23, 1.19) | 0.82  (0.07, 1.77) | .97 | 1.41  (0.47, 2.60) | 1.82  (0.15, 3.98) | .98 |
| Musculoskeletal disorders^d^ | 15.74  (6.70, 28.01) | 11.65  (5.95, 17.94) | .99 | 20.57  (10.07, 33.83) | 18.73  (10.64, 27.90) | .99 |
|  | **EP** | **PT + EP** |  | **EP** | **PT + EP** |  |
|  | n=10 | n=6 |  | n=10 | n=6 |  |
| **Stretcher (n=16)** | **Costs^a^ – Q1 time values** | | ***p*** | **Costs^a^ – Q3 time values** | | ***p*** |
| Average cost of the ED visit | 478.58  (283.12, 733.90) | 579.25  (344.83, 921.39) | .54 | 644.42  (425.98, 927.13) | 713.30  (438.07, 1,084.73) | .71 |
| **ED check-in**^b^ | **139.33**  **(111.60, 185.00)** | **282.35**  **(195.81, 411.38)** | **<.01** | **269.69**  **(231.05, 326.95)** | **393.61**  **(272.61-541.51)** | **.02** |
| ED stay^c^ | **----** | **----** | **---** | **----** | **----** | **---** |
| Imaging | 58.46  (22.46, 98.45) | 30.02  (0.00, 82.18) | .97 | 76.58  (30.53, 126.77) | 39.51  (0.00, 107.07) | .97 |
| Laboratory tests | 15.68  (0.00, 52.25) | 9.23  (0.00, 18.45) | .98 | 18.36  (0.00, 61.21) | 13.71  (0.00, 26.41) | .98 |
| Medication | 12.99  (6.26, 21.82) | 5.91  (2.06, 9.92) | .24 | 24.75  (13.65, 38.39) | 13.12  (4.51, 22.43) | .35 |
| Musculoskeletal disorders^d^ | 16.55  (0.00, 55.17) | 7.94  (0.00, 18.63) | .99 | 19.46  (0.00, 64.88) | 9.56  (0.00, 21.59) | .99 |

EP: emergency physician; PT: physiotherapist; CI: confidence interval; ED: emergency department; Q1: first quartile; Q3: third quartile

^a^ Costs are presented as mean (95% confidence intervals obtained via non-parametric Bootstrap).

^b^ Includes ED arrival (clerk, nurse, nursing assistant), triage reassessment (if needed; nurse), assessment (emergency physician; if needed: physiotherapist, internist, social worker, occupational therapist), reassessment (if needed; emergency physician, physiotherapist), point-of-care ultrasound (if needed; emergency physician).

^c^ Since all the parameters needed to calculate the ED stay cost are fixed (hours billed, patient ratio, ED length of stay), the average costs obtained are the same for all three scenarios.

^d^ Musculoskeletal disorders care processes include supplying and advising on use for walking aids and orthoses (crutches, cane, walker or walking boot), and splinting (plaster cast, Jones splint, thoracobrachial splint, ankle splint or Zimmer splint).
